# Variant-specific pathophysiological mechanisms of *AFF3* differently influence transcriptome profiles

**DOI:** 10.1101/2024.01.14.24301100

**Authors:** Sissy Bassani, Jacqueline Chrast, Giovanna Ambrosini, Norine Voisin, Frédéric Schütz, Alfredo Brusco, Fabio Sirchia, Lydia Turban, Susanna Schubert, Rami Abou Jamra, Jan-Ulrich Schlump, Desiree DeMille, Pinar Bayrak-Toydemir, Gary Rex Nelson, Kristen Nicole Wong, Laura Duncan, Mackenzie Mosera, Christian Gilissen, Lisenka E.L.M. Vissers, Rolph Pfundt, Rogier Kersseboom, Hilde Yttervik, Geir Åsmund Myge Hansen, Marie Falkenberg Smeland, Kameryn M. Butler, Michael J. Lyons, Claudia M.B. Carvalho, Chaofan Zhang, James R. Lupski, Lorraine Potocki, Leticia Flores-Gallegos, Rodrigo Morales-Toquero, Florence Petit, Binnaz Yalcin, Annabelle Tuttle, Houda Zghal Elloumi, Lane Mccormick, Mary Kukolich, Oliver Klaas, Judit Horvath, Marcello Scala, Michele Iacomino, Francesca Operto, Federico Zara, Karin Writzl, Ales Maver, Maria K. Haanpää, Pia Pohjola, Harri Arikka, Christian Iseli, Nicolas Guex, Alexandre Reymond

**Affiliations:** Center for Integrative Genomics, University of Lausanne, Lausanne, Switzerland; Bioinformatics Competence Center, University of Lausanne, Lausanne, Switzerland; Bioinformatics Competence Center, Ecole Polytechnique Fédérale de Lausanne, Lausanne, Switzerland; Biostatistics platform, University of Lausanne, Lausanne, Switzerland; Department of Neurosciences Rita Levi-Montalcini, University of Turin, 10126 Turin, Italy; Medical Genetics Unit, Città della Salute e della Scienza University Hospital, 10126 Turin, Italy; Institute of Human Genetics, University of Leipzig Medical Center, Leipzig, Germany; Department of Pediatrics, Centre for Neuromedicine, Gemeinschaftskrankenhaus Herdecke Gerhard-Kienle-Weg, Herdecke, Germany; Genomics Analysis 396, ARUP Laboratories, Salt Lake City, Utah, USA; Pediatric Neurology, University of Utah School of Medicine, Salt Lake City, Utah, USA; Department of Pediatrics, Medical Center North, Vanderbilt University Medical Center, Nashville, Tennessee, USA; Department of Human Genetics, Research Institute for Medical Innovation, Radboud University Medical Center, Nijmegen, the Netherlands; Center for genetic developmental disorders southwest, Zuidwester, Middelharnis, The Netherlands; Department of Medical Genetics, University Hospital of North Norway, Tromsø, Norway; Department of Pediatric Rehabilitation, University Hospital of North Norway, Tromsø, Norway; Greenwood Genetic Center, Greenwood, South Carolina, USA; Pacific Northwest Research Institute (PNRI), Broadway, Seattle, Washington, USA; Department of Molecular and Human Genetics, Baylor College of Medicine, Houston, Texas, USA; Human Genome Sequencing Center, Baylor College of Medicine, Houston, Texas, USA; Department of Pediatrics, Baylor College of Medicine, Houston, Texas, USA; Texas Children’s Hospital, Houston, Texas, USA; Hospital Ángeles Puebla, Puebla, Mexico; CHU Lille, Clinique de génétique, F-59000 Lille, France; Inserm UMR1231, University of Burgundy, 21000 Dijon, France; GeneDx, Gaithersburg, Maryland, USA; Department of Genetics, Cook Children’s Medical Center, Cook Children’s Health Care System, Fort Worth, Texas, USA; Institute for Human Genetics, University Hospital Muenster, Muenster, Germany; Department of Neurosciences, Rehabilitation, Ophthalmology, Genetics, Maternal and Child Health, University of Genoa, 16132, Genoa, Italy; Medical Genetics Unit, IRCCS Istituto Giannina Gaslini, Genoa, Italy; Child and Adolescent Neuropsychiatry Unit, Department of Medicine, Surgery and Dentistry, University of Salerno, Salerno, Italy; Clinical Institute of Genomic Medicine, University Medical Centre Ljubljana, Ljubljana, Slovenia; Faculty of Medicine, University of Ljubljana, Ljubljana, Slovenia; Department of Genomics, Turku University Hospital, Turku, Finland; University of Turku, Turku, Finland; Department of Pediatric Neurology, Turku University Hospital, Turku, Finland; University of Turku, Turku, Finland

**Keywords:** mesomelic dysplasia, horseshoe kidney, intellectual disability, transcriptome, exome

## Abstract

**Background:** We previously described the KINSSHIP syndrome, an autosomal dominant disorder associated with intellectual disability (ID), mesomelic dysplasia and horseshoe kidney,caused by *de novo* variants in the degron of AFF3. Mouse knock-ins and overexpression in zebrafish provided evidence for a dominant-negative (DN) mode-of-action, wherein an increased level of AFF3 resulted in pathological effects.

**Methods:** Evolutionary constraints suggest that other mode-of-inheritance could be at play. We challenged this hypothesis by screening ID cohorts for individuals with predicted-to-be deleterious variants in *AFF3*. We used both animal and cellular models to assess the deleteriousness of the identified variants.

**Results:** We identified an individual with a KINSSHIP-like phenotype carrying a *de novo* partial duplication of *AFF3* further strengthening the hypothesis that an increased level of AFF3 is pathological. We also detected seventeen individuals displaying a milder syndrome with either heterozygous LoF or biallelic missense variants in *AFF3*. Consistent with semi-dominance, we discovered three patients with homozygous LoF and one compound heterozygote for a LoF and a missense variant, who presented more severe phenotypes than their heterozygous parents. Matching zebrafish knockdowns exhibit neurological defects that could be rescued by expressing human *AFF3* mRNA, confirming their association with the ablation of *aff3*. Conversely, some of the human *AFF3* mRNAs carrying missense variants identified in affected individuals did not complement. Overexpression of mutated *AFF3* mRNAs in zebrafish embryos produced a significant increase of abnormal larvae compared to wild-type overexpression further demonstrating deleteriousness. To further assess the effect of *AFF3* variation, we profiled the transcriptome of fibroblasts from affected individuals and engineered isogenic cells harboring +/+, DN/DN, LoF/+, LoF/LoF or DN/LoF *AFF3* genotypes. The expression of more than a third of the AFF3 bound loci is modified in either the DN/DN or the LoF/LoF lines. While the same pathways are affected, only about one-third of the differentially expressed genes are common to these homozygote datasets, indicating that *AFF3* LoF and DN variants largely modulate transcriptomes differently, e.g. the DNA repair pathway displayed opposite modulation.

**Conclusions:** Our results and the high pleiotropy shown by variation at this locus suggest that minute changes in *AFF3* function are deleterious.

## Background

*AFF3* encodes the ALF Transcription Elongation Factor 3 (MIM*601464), a member of a gene family with four paralogs (*AFF1-4*) in mammals. These nuclear proteins function as transcriptional activators, promoting RNA elongation^1–3^. They share conserved N-terminal (NHD) and C-terminal homology domains (CHD)^4^, an AF4-LAF4-FMR2 (ALF) domain^2,3,5^, which contains the degron motif, a Serine-rich transactivation domain (TAD)^6^, and a nuclear/nucleolar localization sequence (NLS) (Figure 1A). AFF proteins are integral components of transcriptional super elongation complexes (SECs) that include positive transcription elongation factor (P-TEFb)^2,3^. SECs are made of an AFF family member as scaffold, YEATS domain-containing MLLT proteins (myeloid/lymphoid or mixed-lineage leukemia; translocated to), and an ELL (Elongation Factor for RNA Polymerase II) protein^2^. By phosphorylating the C-terminal domain of RNA polymerase II, these complexes regulate the RNA transcription elongation process^3,7^. Distinct combinations of components yield different SECs providing gene target specificity^2,3^. AFF3 regulates the expression of genes involved in mesoderm and ectoderm development, as well as mesenchymal cell proliferation, cell adhesion, angiogenesis, cartilage and lens development and immunoglobulin class switch recombination^8,9^. It was recently linked with the establishment of biological rhythms, e.g. somitogenesis progression and niche switching^10,11^.

**Figure 1.**
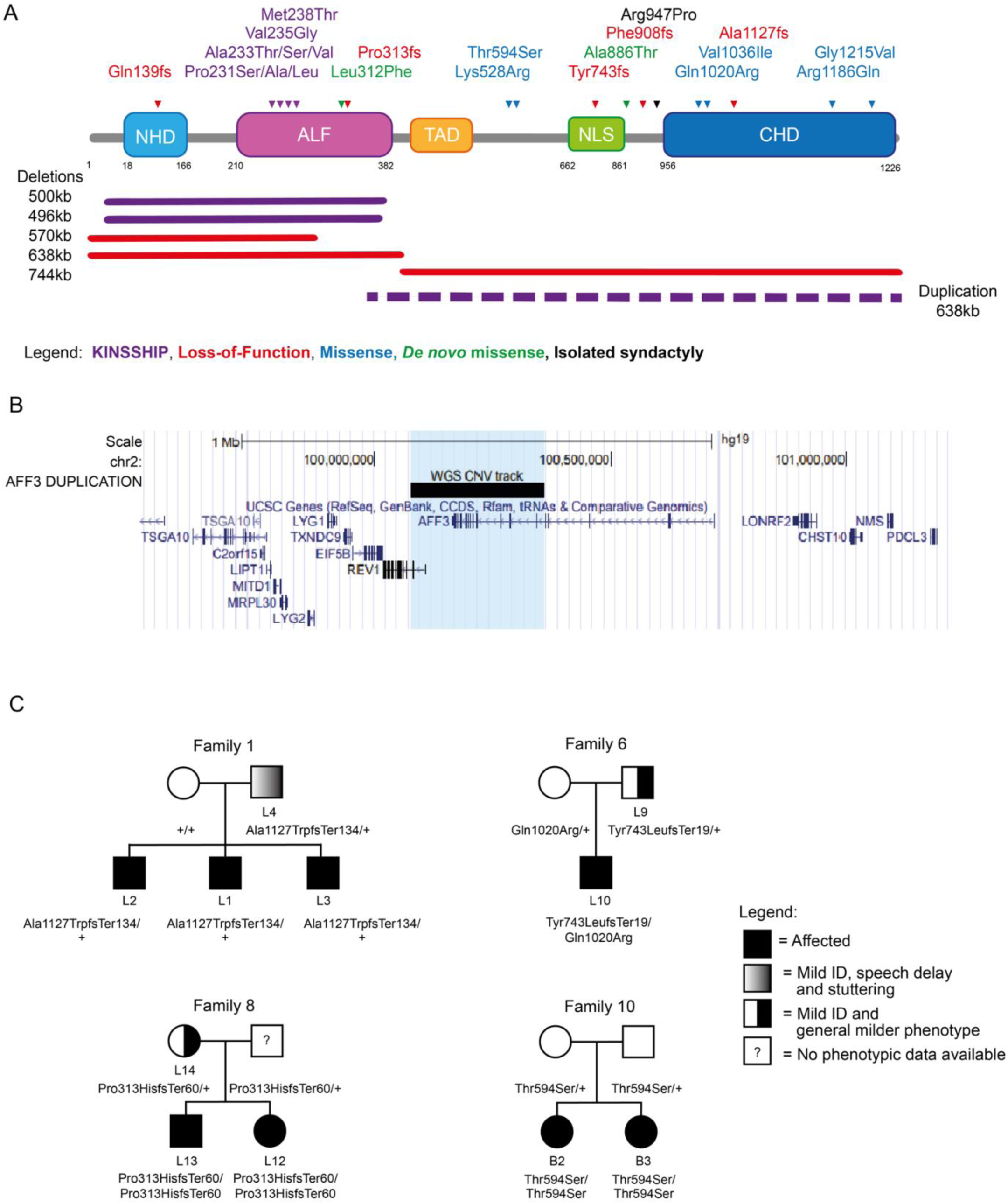
*AFF3* variants. **(A)** Schematic protein structure of AFF3 (NM_002285.3) with its N-terminal homology domain (NHD, cyan), the AF4-LAF4-FMR2 (ALF, pink) domain^2,3,5^ containing the degron motif, a Serine-rich transactivation domain (TAD, yellow)^6^, a bipartite nuclear/nucleolar localization sequence (NLS, green), and the C-terminal homology domain (CHD, blue)^4^ showing positioning of all *AFF3* coding variants mentioned in the text. Missense and truncating variants are shown above, while extents of microdeletions and microduplications are depicted below the structure with continuous and dashed lines, respectively. The variants are color coded: loss-of-function (truncation and deletion) in red, biallelic missense outside the degron in blue and KINSSHIP-associated missense variants, deletion and duplication in purple. The *de novo* missense identified in individuals M1 and M2 are shown in green. While the p.(Arg947Pro) shown in black was shown to segregate with isolated syndactyly^49^., we have also identified it in individuals with no digit abnormalities. **(B)** UCSC genome browser snapshot of the Chr2 99.5 to 101.4 Mb region showing the genes mapping to this interval. The extent of the duplication identified in the DUP1 individual that encompasses exon 10 to exon 24 of *AFF3* and exon 1 to 3 of *REV1* is indicated by the black bar and the light blue shadow. **(C)** Examples of pedigrees of transmitting affected families.

The SEC-L3 complex, which incorporates AFF3, is enriched at imprinted loci, for example at the lncRNA *XIST* locus that initiate X chromosome inactivation^12,13^. AFF3 binds both silent and active chromatin regions to modulate expression of imprinted regions. For example, within the Dlk1-Dio3 interval, it is recruited by ZFP281 to the *Meg3* enhancer region to maintain an active chromatin state through H3K27ac modification and an allele-specific expression^8,14^.

We previously reported the association of *AFF3* alterations with KINSSHIP syndrome^11^. Twenty-one affected individuals allowed delineation of its cardinal characteristics. Such individuals presented with developmental delay/intellectual disability (DD/ID), brain atrophy, epileptic encephalopathy, failure to thrive, horseshoe kidney, a specific mesomelic dysplasia, fibular hypoplasia, scoliosis, hypertrichosis, dysmorphic facial features, gastrointestinal and pulmonary symptoms. This autosomal dominant disease is associated with *de novo* germline missense variants and deletions, as well as mosaic variants, in the conserved degron motif of AFF3^11,15^ (**Figure 1A**). Both mouse knock-ins and overexpression in zebrafish suggested a dominant-negative (DN) mode of action, wherein an increased level of AFF3 resulted in the pathological effects.

According to population metrics presented in GnomAD v2.1.1^16^, *AFF3* is under constraint with a pLI=1 and a pLOEUF=0.221, which suggests that *AFF3* haploinsufficiency could also be deleterious. Consistent with this hypothesis, mosaic CGG trinucleotide-repeat expansions in the promoter of *AFF3* that result in its hypermethylation and silencing, were associated with mild ID, speech and motor delays, seizures, behavioral disturbances, generalized hypotonia, dysmorphic features and congenital anomalies^17,18^.

Here we describe novel *AFF3* genetic alterations associated with an overexpression disease mechanism, as well as the effect of decreased AFF3 function through haploinsufficiency, homozygous truncation and autosomal recessive inheritance. The affected individuals present symptoms that partially overlap those of KINSSHIP.

## Materials and Methods

### Samples and variants identification

Informed consent forms were obtained for all affected individuals or their guardians participating in this study. The current study was approved by the CER (“Commission d’éthique de la recherche”) of the canton of Vaud (Protocol number: CER-VD 2021-01400). This research complies with the principles of the Declaration of Helsinki. The IDs of affected individuals are unknown to anyone outside the research group. Affected individuals underwent genetic counselling and clinical examination followed by exome sequencing as described^19,20^ and/or array comparative genome hybridization, made exception of proband B1 who was sequenced with the Illumina TruSight One Expanded panel covering about one third of the exome. Genome sequencing was performed on the DUP1 individual to characterize the breakpoints.

### Protein model

Alignment of multiple AFF3 orthologous sequences was performed with the Clustal Omega tool ^21,22^. 3D modelling for AFF3 (UniProt: P51826) and SIAH1 (UniProt:Q8IUQ4) interaction was built using the Swiss-Pdb Viewer^23^ as previously described^11^.

### Zebrafish husbandry

Zebrafish (*Danio rerio*, Oregon AB) were maintained at 28.5 °C on a 14:10 h light/dark cycle. Zebrafish are staged by hours (h) or days (d) post fertilization (pf). Adult zebrafish were housed in Active Blue racks (Tecniplast, Buguggiate, Italy) with a maximum of 20 fish per tank. All procedures complied with the European Convention for the Protection of Animals for Experimental and Scientific Purposes (ETS number 123) and the National Institutes of Health guide for the care and use of Laboratory animals. Housing and experiments were approved by the Vaud cantonal authority (authorization VD-H21).

### Zebrafish CRISPR-Cas9 model

We generated founder F0 mutant zebrafish depleted for *aff3* by CRISPR/Cas9 genome editing. Two single synthetic guide RNAs (sgRNAs) targeting the coding sequence in *aff3* exon 6 of both isoforms annotated by Ensembl (Zebrafish GRCz11) (sgRNA_r2 5’- TCCAAAGCAGTACCCAGCCAAGG -3’; sgRNA_r19 5’- GCACCTGAGAATATATACCTTGG -3’) were designed with the CHOPCHOP tool ^24,25^ and ordered from Synthego, Redwood City, CA, USA. A total of 1nl of a cocktail containing 50 ng/μl of gRNA_r2, 50 ng/μl of gRNA_r19 and 200 ng/μl of TrueCut^TM^Cas9 v2 (Invitrogen) was injected into one to two-cell stage embryos. In mock-injected larvae, the Cas9 was replaced by the same volume of water. KCl (200 mM) was added to increase efficiency. To determine the CRISPR-Cas9 targeting efficiency of each sgRNA in 5dpf founder (F0) mutants, a mismatch detection assay using T7 endonuclease 1 (New England Biolabs, Ipswich, MA, United States) was performed. Briefly, DNA was extracted, and PCR amplified with primers flanking the sgRNAs target site (5’- TCCAAAGCAGTACCCAGCCAAGGTATATATTCTCAGGTGC -3’). PCR products were denatured, reannealed, and incubated with T7 for 15 minutes at 37°C. The reaction was stopped by adding 1.5 μl of 0.25 M EDTA. The products were then separated on 2% agarose gel to determine rearrangements at the targeted site.

### Locomotion assays

At 72 hpf, the escape response test was performed to evaluate the swimming ability of the fish upon a slight touch stimulation. The motion of every larva was examined and scored as « normal swimming », « pause », « looping swimming », « pinwheel swimming » or « motionless » due to malformations. At 5 dpf, we analyzed spontaneous zebrafish motility using the Zebrabox^®^ recording system (Viewpoint, Lissieu, France) equipped with infrared illumination for imaging in the dark. Locomotion was recorded for each larva on a 96-well plate for 30 minutes (15-minute adaptation phase in the light followed by a 15-minute phase in the dark) and presented as slow (3-6 mm/s) and high velocities (>6 mm/s)^26^. The velocity of the fish was tracked with the Viewpoint software and experiments were performed at minimum three times. The resulting data were pooled together for statistical analysis. Fisher’s exact test or one-way ANOVA analysis were performed based on the data with Prism10.

### Immunofluorescence

PTU (1-phenyl 2-thiourea - 75μM) treatment was used on 24hpf zebrafish to prevent pigmentation. At appropriate developmental stages, embryos were dechorionated and euthanized with 0.0168% tricaine (Sigma-Aldrich) and immediately fixed in 4% PFA for 1h at RT. Permeabilization of larvae was achieved with 1X phosphate saline buffer (PBS), 0.5% Triton X-100, for 90 min at RT and subsequently in 1X PBS, 1%Triton X-100, for 2h at RT on a slow shaker. Embryos were then incubated in blocking buffer (1% BSA in 1X PBS) for 1h at RT and incubated in primary antibodies, mouse anti-synaptotagmin 2 (Znp-1, diluted 1:100 in blocking solution – DSHB, Iowa City, IA, United States) or mouse anti-islet 1 and 2 (39.4D5, diluted 1:100 in blocking solution – DSHB, Iowa City, IA, United States), overnight at 4°C on a slow shaker. After 3 washes in 1X PBS, the embryos were incubated with a secondary antibody, Alexa Fluor™ 488 conjugated (diluted 1:500 in blocking solution, Invitrogen), overnight at 4°C. Nuclei were stained with DAPI (diluted 1:8000, Sigma-Aldrich) for 5/10 min at RT. After washing in PBS, zebrafish larvae were mounted onto microscopic slides with Mowiol 4-88 (Sigma-Aldrich). Imaging was performed using LSM880 airyscan confocal microscope (Carl Zeiss). Evaluation of motor neurons’ structure and hindbrain spinal cord projecting neurons’ development was performed^27^.

### Morphological analyses

Images of 5dpf zebrafish were acquired with a Leica microscope (M165 FC) and Leica CMOS camera (IC90E, Leica Camera AG, Wetz-lar, Germany) for morphological inspection. Inter-ocular distance and head width were quantified using the Fiji software^27^. Fisher’s exact test or one-way ANOVA analysis were performed with Prism10.

### Staining of cartilaginous structure

At 5dpf, embryos were euthanized with 0.0168% tricaine (Sigma-Aldrich) and fixed overnight in 4% PFA at RT. Fixed embryos were washed four times with 1X PBS and 0.1% Tween-20 (PBST) and bleached with 30% hydrogen peroxide for 2h at RT. After three wash cycles with PBST, specimens were transferred into an Alcian Blue solution (1% concentrated hydrochloric acid, 70% ethanol, 0.1% Alcian blue) and stained overnight at 4 °C. Embryos were rinsed a few times with acidic ethanol (5% concentrated hydrochloric acid, 70% ethanol, HCl-EtOH) and incubated in acidic ethanol for 20min at RT on a slow shaker. Specimens were then re-hydrated as follows: (i) 5/10min at RT in 1mL of 75% HCl-EtOH / 25% H_2_O_d_; (ii) 5/10min at RT in 1mL of 50% HCl-EtOH / 50% H_2_O_d_; (iii) 5/10min at RT in 1mL of 25% HCl-EtOH / 75% H_2_O_d_ and (iv) 5/10min at RT in 1mL of 100% H_2_O_d_. Specimens were stored in 1mL of 50% Glycerol and 50% (1%) KOH or kept in 100% Glycerol for extended storage. Stained embryos were positioned in 50% Glycerol and 50% (1%) KOH solution in a Petri dish. The head was photographed in a ventral–dorsal and a lateral view using a stereo microscope (Motic SMZ-171) with the Motic Image Plus software (version 3.0).

### Overexpression analysis in zebrafish

Tagged human *AFF3* wild-type mRNA (GenBank: NM_002285.3) was cloned into pEZ-M13 vector^11^. The variants of interest, i.e. the two KINSSHIP variants Val235Gly and Ala233Thr and the three newly-identified missense variants Gln179Glu, Lys528Arg and Thr594Ser, were engineered using the QuikChange II XL Site-Directed Mutagenesis Kit following the manufacturer’s instructions (Agilent Technologies). Positive clones were confirmed by Sanger sequencing. *AFF3* mRNA was transcribed from the linearized vector pEZ-M13+AFF3-FLAG Wt^11^ or containing each of the studied variants using the mMESSAGE mMACHINE T7 transcription kit (Ambion) and purified using RNeasy Mini Kit (Qiagen) following the manufacturers’ instructions. The injection mix consisted of the mRNAs at three different concentrations (180 ng, 360 ng and 720 ng) diluted in RNAse-free water. 1nl of each diluted mRNA was injected inside the yolk, below the cell, in AB wild-type zebrafish embryos at the one to two-cell stage. Distilled water was injected as vehicle control in a similar volume. Depending on RNA amounts, experiments were repeated twice or three times.

### Phenotypic Rescue in zebrafish

We engineered F0 zebrafish depleted for *aff3* by CRISPR/Cas9 genome editing and expressing the human *AFF3* mRNA of interest. The rescue experiment was conducted by evaluating the spontaneous zebrafish motility in the dark using the Zebrabox^®^ recording system (Viewpoint, Lissieu, France). The injection mix consisted of sgRNAs/Cas9 complex and human *AFF3* mRNA Wt (for the phenotypic rescue) or the human AFF3 mRNA carrying each variant of interest. Different concentrations of human AFF3 mRNA Wt (25 ng, 50 ng, 75 ng, 100 ng, 150 ng, 200 ng) were tested to reach the phenotypic rescue. Phenotypic rescue by the variants was assessed by injecting 1nl of mix containing sgRNAs/Cas9 complex + 75ng (75 pg/μl) of each mRNA into one or two-cell stage embryos. The sgRNAs were injected alone in mock-injected larvae with Cas9 replaced by the same volume of water. The GraphPad Prism software (version 10.0) was used to perform statistical analysis of the data. A one-way ANOVA test was adopted to determine differences between experimental groups. Experiments were performed seven times.

### HEK293T isogenic cell lines

HEK293T cells were used to engineer *AFF3* knock-ins (KINSSHIP) and knockouts (LoF) cell lines by CRISPR/Cas9 genome editing. Four guides were used to create the LoF lines: one targeting the coding sequence of exon 6, designed with the Thermo Fisher Scientific tool, and three targeting exon 5 with the Gene Knockout Kit v2 of Synthego. To engineer the KINSSHIP lines, one sgRNA targeting the coding sequence in exon 6 designed using the Thermo Fisher Scientific tool, was combined with a DNA donor template to knock-in the Ala233Thr variant. The sgRNAs, DNA donor template and corresponding sequencing primer pairs were ordered at Invitrogen, Synthego, or Sigma-Aldrich. The cocktail to induce AFF3 knock-out contained 7.2μg of the four combined sgRNAs and 36.2 μg of TrueCut^TM^Cas9 v2 protein (Invitrogen). In the KINSSHIP model, 7.2μg of the sgRNA by Thermo Fisher Scientific, combined with 36.2 μg of TrueCut^TM^Cas9 v2 protein and 14.5 μg ds DNA donor, were used. Each mix was transfected using the Lipofectamine™ CRISPRMAX™ Cas9 Transfection Reagent Kit (Invitrogen) on 10 cm HEK293T cell plates according to the manufacturer’s protocol. 48h after transfection, cells were collected, resuspended post-counting, and diluted at a density of 8 cells/ml. 100 μl of this resuspension was transferred to each well of a 96-well plate. At the desired cell confluency, clones were screened with the QIAprep&amp CRISPR kit (QIAstock, QIAGEN, AG). Variants were confirmed by Sanger sequencing. Heterozygotes were further confirmed by cloning and sequencing of both alleles. We engineered five biallelic LoF HEK293T lines (LoF/LoF) with different combinations of variants (lines No.20 and 98: stop-gain/stop-gain; No.15: stop-gain/20bp deletion; No.4: 4bp deletion/114bp deletion; No.216: 94bp deletion/94bp deletion), one heterozygous LoF stop-gain/+ line (No.1), two homozygous Ala233Thr/Ala233Thr KINSSHIP/KINSSHIP lines (No.54 and 90) and two compound heterozygous KINSSHIP and LoF lines (No.51 and 86: Ala233Thr/stop-gain). These ten lines and three unmutated HEK293T lines were grown simultaneously in biological triplicate before RNA extraction with RNeasy Mini Kit (QIAstock, QIAGEN AG). The nomenclature of the engineered variants is:

A. *Stop gain (through A insertion)*: GRCh37:2:100623265:A:AT, NM_002285.3:c.701dup, NP_002276.2:p.(Tyr234Ter) NC_000002.11:g.100623266dup
B. *4bp del*: GRCh37:2:100623262:CACAT:C, NM_002285.3:c.701_704del, NP_002276.2:p.(Tyr234Ter) NC_000002.11:g.100623265_100623268del
C. *20bp del*: GRCh37:2:100623247:GCCGTCCATTGGCCTCACATA:G, NM_002285.3:c.700_719del, NP_002276.2:p.(Tyr234ProfsTer6) NC_000002.11:g.100623249_100623268del
D. *94bp del*: GRCh37:2:100623727:ACGAGGGCTGGTTCTGGGCTCTTGAATCTGCAACAAAATGTTCATCGATCTTGTTCACAGGAGTCTGAGGAACCCCAGGTTTGGGAACTCCAACG:A, NM_002285.3:c.276_369del, NP_002276.2:p.(Val93LeufsTer97) NC_000002.11:g.100623728_100623821del
E. *114bp del*: GRCh37:2:100623258:GCCTCACATACGCGGTCGGTTTCTGCTGGACCAGGCTGGGTTTTGAAGCTAGGGATGGAGGAAAGTTCTGAACACAGTGTCCGCTGCTGCTGTGCTTGGCCGCCATGGCAGGTGGC:G, NM_002285.3:c.594_708del, NP_002276.2:p.(Arg198SerfsTer16) NC_000002.11:g.100623260_100623374del
F. *Ala233Thr KINSSHIP variant*: GRCh37:2:100623268:CGC:GGT, NM_002285.3:c.697_699delinsACC, NP_002276.2:p.(Ala233Thr) NC_000002.11:g.100623268_100623270delinsGGT

### Fibroblasts

Fibroblast cells from two patients’ skin biopsies and three healthy age-matched control individuals (2-16 years of age) were grown simultaneously. At the desired cell confluency, RNA was extracted with RNeasy Mini Kit (QIAstock, QIAGEN AG).

### Transcriptome profiling

RNA quality was assessed on a Fragment Analyzer (Agilent Technologies). The RNAs had RQNs between 9.0 and 10.0. RNA-seq libraries were prepared from 500 ng of total RNA with the Illumina TruSeq Stranded mRNA reagents (Illumina) using a unique dual indexing strategy, and following the official protocol automated on a Sciclone liquid handling robot (PerkinElmer). Libraries were quantified by a fluorometric method (QubIT, Life Technologies) and their quality assessed on a Fragment Analyzer (Agilent Technologies). Sequencing was performed on an Illumina NovaSeq 6000 for 100 cycles single read. Sequencing data were demultiplexed using the bcl2fastq2 Conversion Software (version 2.20, Illumina). We profiled transcriptomes with a minimum of 17.9 and 52.9 million mapped reads for HEK293T and fibroblasts, respectively. The HEK293T and fibroblast reads are deposited in GEO under accession GSE241621 (token otezqcsslbctbyx) and GSE246554 (token wxibcyqatdyphkl), respectively Raw reads were aligned to the human (hg38) genome using STAR (2.7.10b), the exact parameters are: STAR^28^ --runMode alignReads --twopassMode Basic --outSAMtype BAM SortedByCoordinate --outSAMattributes All --readFilesCommand “gzip -dc” --quantMode GeneCounts. Gene counts were generated using FeatureCounts^29^ and differential expression analysis was performed with the DESeq2 (v.1.36.0)^30^ package from Bioconductor (v3.15) ^31^. Genes were considered differentially expressed based on an adjusted p-value cutoff of <0.05. Pathway enrichment analysis was carried out using clusterProfiler (v.4.4.4)^32,33^ from Bioconductor using the enricher function. GSEA^34^ analysis was carried out using the GSEA function in ClusterProfiler, and the following annotated gene sets from MSigDB v6.2^35^: the Hallmark gene set^36^. For comparison with ChIP-seq studies in human HEK293T^13^ and ES mouse cell lines^14^, external sequencing data in bigWig format were acquired from GEO. UCSC bigWig files were created at 1bp resolution and normalized to total alignable reads (reads-per-million). Peak detection was performed with MACS v3.0^37,38^ using the bdgpeakcall function (with cut-offs 0.4 and 0.6 respectively). The AFF3 peak regions in mice were lifted over to the hg38 human genome assembly. The peak regions were annotated in R using the ChIPseeker^38^ package, in particular the “annotate Peak” function.

## Results

### KINSSHIP probands

Through data aggregation, we identified three more KINSSHIP individuals (K22-K24), two of whom (K22-K23) harbor previously unreported *de novo* variants (**Figure 1A**; **Table S1**). Two more individuals (K25-K26) carrying previously detected variants^11^ are described in reference^39^. The p.(Pro231Ser), p.(Ala233Ser), p.(Ala233Thr), and p.(Met238Thr) variants of K22-K26 fall within the nine amino acids long 230-KPTAYVRPM-238 degron motif and further expand the number of its residues whose modification is associated with KINSSHIP (i.e. Pro231, Ala233, Val235 and Met238; numbering according to NM_002285.3 throughout) (**Figure 1A**). Pathogenicity of the previously undescribed missense variants is supported by the 3D representation of the encoded degron peptide (**Figure S1**). Whereas changes at Pro231 were previously suggested to affect the backbone kink conferred by this conserved residue^11^, the Met238 sidechain is pointing outward, forward-facing the Ser154 sidechain of a SIAH ubiquitin ligase loop. Modeling suggests that variants at this position should slightly alter binding, predicting a less severe phenotype. Consistent with this hypothesis, proband K22 presented with mild DD, mild speech impairment, facial dysmorphisms, skeletal malformations, mild hypertrichosis, and mild hypotonia, a phenotype milder than that of typical KINSSHIP individuals with variants of residues that dock in the ubiquitin ligase binding pocket^11^.

### Duplication proband

Data aggregation also enabled ascertainment of an individual with a KINSSHIP-like phenotype carrying a *de novo* partial duplication of *AFF3* further strengthening the hypothesis that an increased level of AFF3 is pathological. This DUP1 proband presented with severe failure to thrive with postnatal onset, severe DD with poor eye contact, poor head control, inability to sit and speak, epilepsy, corpus callosum hypoplasia, facial dysmorphism, hypertrichosis, hypotonia, hip, knee, ankle and wrist flexion contractures and severe scoliosis. Whole genome sequencing revealed a tandem duplication of the interval encompassing exon 10 to exon 24 of *AFF3* encoding part of the ALF domain, the TAD, NLS and CHD domains and exon 1 to 3 (up to intron 3-4) of the same orientation ubiquitous *REV1* (chr2:g.100,077,649_100,359,928dup (hg19), NC_000002.11:g.100,077,649_100,359,928dup) (**Figure 1A-B**). The expression of the partially duplicated copy of *AFF3* is then under the control of the *REV1* promoter, which could result in the expression of a degron-less AFF3, a hypothesis that we could not further test directly due to lack of available sample.

### Heterozygous LoF and biallelic probands

To further challenge the hypothesis that diminished expression of *AFF3* is deleterious, we searched for individuals with loss-of-function (LoF) variants in *AFF3*. Using data aggregation of multiple laboratories and clinical centers, e.g. GeneMatcher^40^ and DECIPHER^41^, we identified ten affected individuals with mono-allelic (individuals L1-L9 and L14) and three (L11-L13) with biallelic *AFF3* truncating variants, as well as a proband compound heterozygous for a LoF and a rare missense variant (L10; **Figure 1A and 1C**; **Table S2**). Of note, one of the affected individuals described in reference^18^ was a compound heterozygote for a CGG expansion and a deletion of the *AFF3* promoter. The identified truncating variants are not described in GnomAD (v2.1.1) (**Table S2**). Consistent with the deleteriousness of diminished or absence of *AFF3* expression, these fourteen individuals shared common phenotypes such as global DD/ID (11 out of 11), abnormal corpus callosum (4/6), speech impairment (10/11), muscle disorders/hypotonia (7/9), facial dysmorphisms (6/7), mild cranial dysmorphisms (3/8) and skeletal defects (4/7). To respect medrxiv guidelines and keep affected individuals unidentifiable the list of all symptoms is not included in this preprint, but can is available upon request to the corresponding author. This group counts 10 males and 4 females suggestive of a higher “male susceptibility” (*p*=0.089)^42^. The siblings L12 and L13, who are homozygous for a truncating variant, and L10, who is compound heterozygote for a LoF variant and missense p.(Gln1020Arg) present a more severe phenotype than their parents who are heterozygotes for the LoF variant (families 6 and 8; **Figure 1A and 1C**; **Table S2**). Our search also identified four affected individuals with biallelic homozygous (B1-B3) or compound heterozygous (B7) missense variants in *AFF3* (**Figure 1A and 1C**; **Table S2**). A consanguineous family with three affected individuals was previously described in reference^43^ (B4-B6). These variants are either not described in GnomAD and/or affect the expression of *AFF3* (see below and **Table S2**). They present overlapping symptoms such as DD/ID (6/7) and ADHD (Attention Deficit Hyperactivity Disorder) (2/3), epileptic encephalopathy or abnormal sleep EEG (Electroencephalography) (3/4), short/no attention span (3/4), speech impairment (3/4), heart defects (2/4), and vision impairment (2/4), and other defects. These two cohorts showed a milder phenotype than KINSSHIP probands, suggesting they might represent a new syndrome.

*In silico* modeling of most of the identified missense variants is hampered by the lack of reliable AFFs structural information with the exception of the CHD that is important for dimerization and the ALF that contains the degron and the ELL-binding domains (ELLbow, see below)^44,45^. The p.(Gln1020Arg), p.(Val1036Ile), p.(Arg1186Gln) and p.(Gly1215Val) variants fall within the CHD (**Figure 1A**). A bulky sidechain at position 1215 will collide with Leu1063 and/or Leu1192. Likewise, Val1036 is optimally surrounded by the hydrophobic sidechains of Leu1068, Leu1071, Tyr1072 and Met1075, and cannot accommodate the bulkier p.(Val1036Ile) variant without affecting local packing. Gln1020 and Arg1186 are located at the domain surface, where changes in the local charge might affect binding specificity.Lastly, it is possible that some missense variants outside of the degron are linked to an autosomal dominant disease, as we identified an individual carrying a *de novo* p.(Ala886Thr) variant presenting with DD, speech impairment and ASD (Autism Spectrum Disorder) symptoms and as an individual with a *de novo* p.(Leu312Phe) variant presenting with DD was described in reference^39^ (M1-M2, **Figure 1A**, **Table S2**). The p.(Leu312Phe) variant maps to the ELLbow and 3D models suggest that four of the five possible Phe rotamers will severely clash with either the AFF3 Phe329 or the ELL2 His618 residue.

### Animal models

To further assess if the diminished expression of *AFF3* was deleterious to organismal phenotypes we knocked-down (KD) *aff3*, the zebrafish ortholog of *AFF3*, using CRISPR-Cas9 genome editing. We used two single guide RNAs targeting exon 6 each providing more than 90% efficiency. At 5 days post fertilization (dpf), we observed malformations in 10% of KD larvae, including incomplete eye pigmentation, altered head structure, lateral belly edema, pericardial edema and skeletomuscular dysmorphology (**Figure 2A**). Staining of the cartilaginous cranial structure revealed malformations in 75% of KD larvae (**Figure 2B**). inter-ocular distance (IOD) and head width (HW) were significantly decreased in *aff3* KD compared to uninjected (Un) (*p*=0.011 IOD and *p*=0.001 HW) and mock (M) injected larvae (*p*=0.041 IOD; *p*=0.004 HW) (**Figure 2C-E**). The escape response test upon a tactile stimulus performed at 3 dpf showed that while none of the mock-injected zebrafish showed perturbed escape responses, about a third of the *aff3* KD larvae were affected (*p*<0.0001). Whereas the majority presented either looping (22.5%) or pinwheel swimming (4.7%), behaviors linked to neurological and mechano-sensory system impairment^26,46,47^ and 5.6% were motionless due to extensive malformations (**Figure 2F**). At 5dpf, the locomotion ability was quantitatively evaluated with an automated tracking device. The *aff3* KD larvae showed a statistically significant decrease in global swimming velocity in the dark compared to Un (*p*<0.0001) and M larvae (*p*=0.0025) (**Figure 2G**). As such hypo-locomotion is often associated with neuromotor deficits and akinesia^46^ we immunostained *aff3* KD larvae hindbrain and motoneurons. Hindbrain Mauthner cells^47^ presented a general developmental delay, and while normal in growth and architecture, motoneurons were disorganized in deformed larvae (**Figure S2**). Similarly, we previously showed that the orthologous mouse knockouts, *Aff3*^+/-^ and *Aff3*^-/-^ C57BL/6N, exhibit skeletal defects, an abnormal skull shape, kidney defects and neurological dysfunction^11,48^. Homozygous *Aff3*^-/-^ exhibited significantly enlarged lateral ventricles and decreased corpus callosum size, when compared with both wild-type and *Aff3^+/-^* males^11,48^. Our zebrafish and mouse results support the contention of causation for *AFF3* LoF variants.

**Figure 2.**
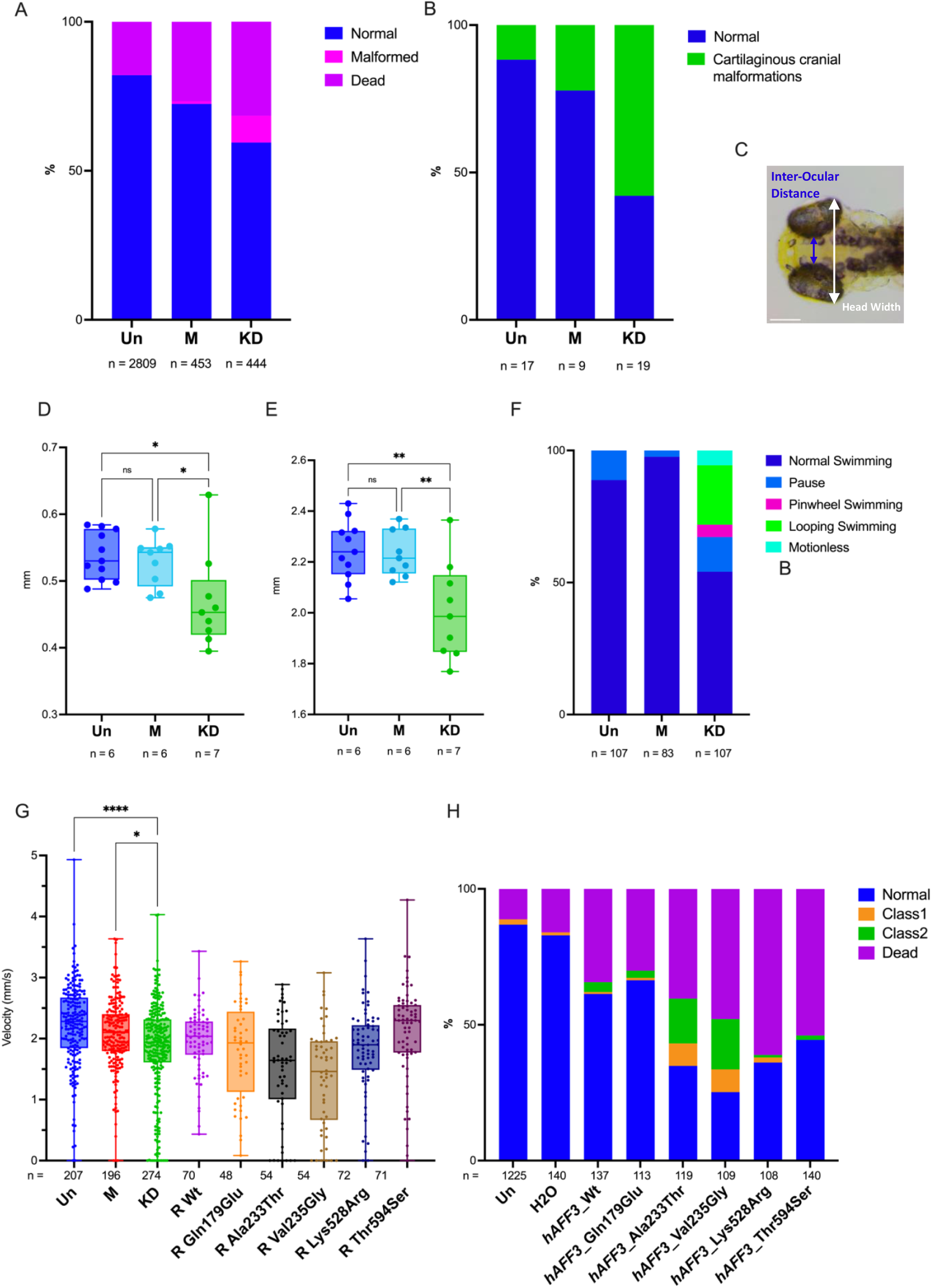
*aff3* knocked-down zebrafish larvae display altered behavior and morphological anomalies. The conditions analyzed are the following: Untreated (Un), Mock-injected (M), and *aff3* Knock-Down (KD). **(A)** Proportions of normal and developmentally defective 5 dpf embryos. In 10% of *aff3* KD zebrafish we identified several morphological anomalies such as: head malformations, belly and heart edema, skeleton-muscular dysmorphologies, and alteration of eye pigmentation. (**B)** Alcian-blue staining at 5 dpf revealed jaw malformation in 57% of *aff3* KD zebrafish. **(C)** Visualization of morphological Inter-Ocular Distance (IOD) and Head Width (HW) measurements from dorsoventral zebrafish image. Quantification of IOD **(D)** and HW **(E)** indicates a significative decrease in IOD and HW in *aff3* KD larvae; *p*\*<0.04; *p*\*\* < 0.0049. **(F)** Touch test response assay at 3 dpf. Upon a touch stimulus, we classified the larvae swimming behavior in: «normal swimming», «pause», «looping swimming», «pinwheel swimming» or «motionless» due to malformations. **(G)** Swimming global velocity analysis at 5dpf in the dark of Un, M, and *aff3* KD and *aff3* KD co-injected with human *AFF3* (*hAFF3*) mRNA wild-type (Wt) or harboring the indicated missense variant, i.e. the KINSSHIP variants Ala233Thr and Val235Gly or the biallelic variants identified in this report in a healthy (Gln179Glu) or affected individuals (Lys528Arg and Thr592Ser); *p*\*\*<0.0025; *p*\*\*\*\* < 0.0001. **(H)** Proportions of normal and developmentally defective 5 dpf embryos uninjected (Un), injected with water as control (H_2_0) or with 360ng of h*AFF3* mRNA Wt or the indicated missense variant. Larvae were cataloged as described: (i) normal phenotype, (ii) Class 1 with skeletomuscular dysmorphology and/or small dimension, (iii) Class 2 with a more severe phenotype including at least three of the following characteristics: skeletomuscular dysmorphology, small dimensions, head malformations, eyes’ alteration, pericardial edema, and lateral belly edema or (iv) dead. Injections of 180ng and 720ng of h*AFF3* mRNA showed similar results.

### Assessing variants

We previously showed that overexpression in zebrafish embryos of human *AFF3* leads to a dose-dependent increase of developmental anomalies^11^, a phenotype that was further exacerbated upon overexpression of the p.(Ala233Thr) KINSSHIP isoform^49^. To assess the pathogenicity of the missense variants identified in the biallelic individuals, we injected zebrafish with human *AFF3* mRNA wild-type (Wt), two selected missense variants present in homozygous state in probands B1 and B2 and his affected sister B3 and mapping outside of crystalized domains (Lys528Arg and Thr594Ser), two KINSSHIP variants (Ala233Thr and Val235Gly) and as control Gln179Glu (Chr2 (GRCh37) g.100623432:G>C, c.535C>G), a variant not described in GnomAD, we identified in homozygosity in a healthy individual. The resulting 5dpf larvae were cataloged as described^49^: (i) normal phenotype, (ii) Class 1 with skeletomuscular dysmorphology and small dimension, (iii) Class 2 with a more severe phenotype including at least three of skeletomuscular dysmorphology, small dimensions, head malformations, eyes’ alteration, pericardial edema, and lateral belly edema or (iv) deceased. Consistent with previously published observations, accumulation of *AFF3* Wt mRNA significantly increased the number of larvae with debilitating traits (*p*=0.0002). Compared to *AFF3* Wt mRNA accumulation, both Ala233Thr and Val235Gly isoforms led to a further significant increase in the number of malformed larvae and mortality at all doses (*p*<0.0001) (**Figure 2H**). Overexpression of the two missense variants identified in proband B1 and B2 similarly caused higher malformations and mortality rates than overexpression of AFF3 Wt (Lys528Arg p<0.0001 and Thr594Ser p=0.0018) albeit not at the rate of the KINSSHIP variants. On the contrary, the control variant p.(Gln179Glu) had an effect similar to that of Wt overexpression (p=0.7; **Figure 2H**).

These results suggest that like truncating variants, at least some of the missense variants identified in the affected individuals could be causative. To challenge this hypothesis further, we performed a phenotypic rescue experiment^50^. As described above, depletion of *aff3* in 5 dpf zebrafish larvae resulted in decreased swimming velocity. That decrease could be rescued by co-injection of human *AFF3* Wt mRNA demonstrating first that human AFF3 can compensate for the loss of its zebrafish ortholog and second that ablation of *aff3* activity was causative of the phenotype (**Figure 2G**). Consistent with the detrimental effect of the overexpression of the KINSSHIP variants, we observed an even lower average velocity upon co-injection of Val235Gly and Ala233Thr mRNAs (both p<0.001 compared to injection of Wt; both *p*<0.0001 compared to M). While co-injection of Lys528Arg mRNA did not rescue *aff3* ablation, co-injection of Thr594Ser mRNA resulted in an “over-rescue” with injected larvae presenting an increased velocity compared to mock (**Figure 2G**). The control variant p.(Gln179Glu) had an intermediate effect halfway between Wt and Lys528Arg mRNA injections. Together these results suggest that these missense variants impact the activity of AFF3 and that biallelic *AFF3* variants could be associated with an autosomal recessive disease. Consistent with the latter hypothesis neither homozygous nor compound heterozygous classified as ‘weak missense variant or worse’, i.e. with a MAF ≤1% and REVEL score ≥0.644^51^ were identified in GnomAD v2.1.1.

### Transcriptome profiling

To compare the transcriptional consequences of AFF3 loss and overexpression, we used CRISPR-Cas9 genome editing to engineer multiple DN and LoF variants in an isogenic cell model, the human embryonic kidney 293T line. HEK293T was chosen (i) as KINSSHIP individuals often present with a horseshoe kidney^50^, (ii) as *AFF3* is expressed in this cell line, and (iii) as in this line both transcriptome profiles of *AFFs* shRNAs knockdowns^3^ and (iv) ChiP-seq of AFF3 have been published^13^. We engineered five biallelic LoF HEK293T lines (LoF/LoF) with different combinations of variants (lines No.20 and 98: stop-gain/stop-gain; No.15: stop-gain/20bp deletion; No.4: 4bp deletion/114bp deletion; No.216: 94bp deletion/94bp deletion), one heterozygous LoF stop-gain/+ line (No.1), two homozygous Ala233Thr/Ala233Thr KINSSHIP/KINSSHIP lines (No.54 and 90) and two compound heterozygous KINSSHIP and LoF lines (No.51 and 86: Ala233Thr/stop-gain). We profiled the transcriptomes of three biological replicates of each of these lines by RNA-sequencing and compared them to those of three biological replicates of three wild type lines (Wt1, Wt2 and Wt4), for a total of 39 profiles. While *AFF3* mRNA levels are significantly decreased in the five biallelic LoF/LoF lines (*padj*=3.5E-53), the Ala233Thr/Ala233Thr KINSSHIP/KINSSHIP and the LoF/+ lines present *AFF3* transcript levels comparable and intermediate (*padj*=0.011) to that found in control Wt lines, respectively (**Figure S3**). The similar level of expression of *AFF3* in the wild type and the Ala233Thr/Ala233Thr lines suggest that there is no negative feedback loop to balance the amount of AFF3. We first compared the transcriptome of homozygous LoF/LoF and Ala233Thr/Ala233Thr lines to that of +/+ lines and identified 3,553 and 4,177 differentially expressed genes (DEG) at an adjusted p-value threshold of 0.05, respectively (**Figure 3A-B**, **Table S3-S4**). We observed an overlap of 23% of DEGs with previous transcriptome profiling of HEK293T cells in which *AFF3* was knocked down with shRNAs^3^.

**Figure 3.**
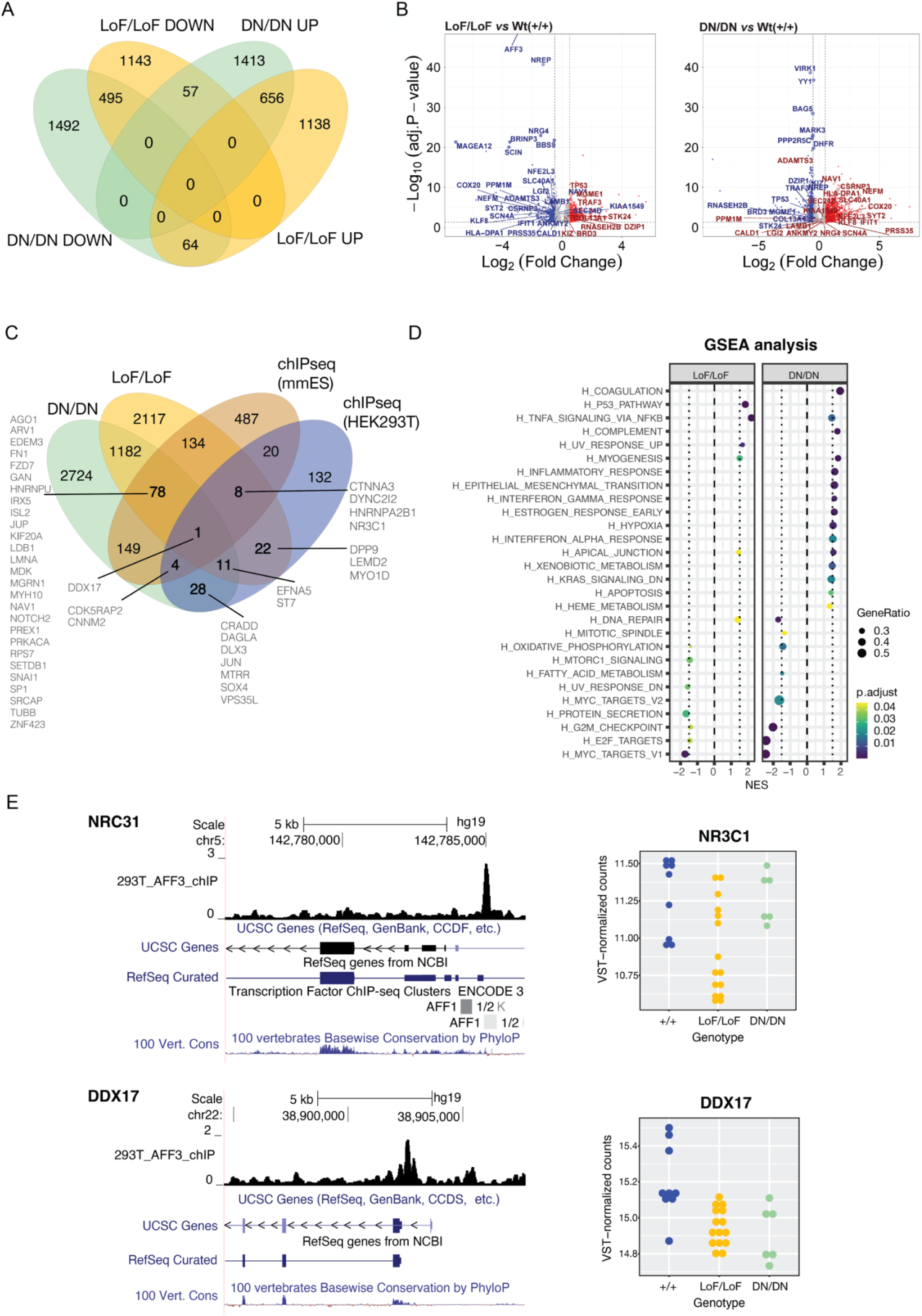
Transcriptome profiles of engineered isogenic HEK293T cells. **(A)** Four-way Venn diagram of differentially expressed genes (DEGs) in biallelic loss-of function (LoF/LoF) *AFF3* lines and biallelic dominant negative (DN/DN) *AFF3* KINSSHIP lines upon comparison with unmutated wildtype lines. DEGs counts are stratified in genes up- (UP) and down-regulated (DOWN). **(B)** Volcano Plots of DEGs in biallelic loss-of function (LoF/LoF) *AFF3* lines (left panel) and biallelic dominant negative (DN/DN) AFF3 KINSSHIP lines (right panel) upon comparison with unmutated wildtype lines. The top 30 most significant DEGs in LoF/LoF that are dysregulated in an opposite manner in DN/DN are indicated, together with some of the most differentially expressed genes (-log10(Padj)>20 and abs(log2FoldChange) > 0.5). **(C)** Gene Set Enrichment Analysis (GSEA) for hallmark pathways of DEGs in biallelic loss-of function (LoF/LoF) *AFF3* lines (left panel) and biallelic dominant negative (DN/DN) *AFF3* KINSSHIP lines (right panel) upon comparison with unmutated wildtype lines. **(D)** Four-way Venn diagram of differentially expressed genes (DEGs) in biallelic loss-of function (LoF/LoF) *AFF3* lines and biallelic dominant negative (DN/DN) *AFF3* KINSSHIP lines upon comparison with unmutated wildtype lines and AFF3 ChIP-seq peaks identified in HEK293T cells (HEK293T) and in *Mus musculus* ES cells (mmES). DEGs bound by AFF3 discussed in the text are indicated. **(E)** Examples of DEGs *NRC31* (top) and *DDX17* (bottom) loci bound by AFF3. UCSC genome browser snapshot showing from to top to bottom AFF3 ChIP-seq HEK293T results, UCSC and REFSeq curated gene structure and vertebrate PhyloP conservation scores (left panels). Expression level of *NRC31* (top) and *DDX17* (bottom) in +/+ (blue), LoF/LoF (yellow) and DN/DN (green) HEK293T engineered lines (right panels).

The LoF/LoF and KINSSHIP/KINSSHIP (DN/DN) lines present with significant repression of genes involved in the G2-M transition, oxidative phosphorylation, targets of E2F and MYC-related genes, most markedly in the DN model. Both lines showed an upregulation of the inflammatory response (e.g TNFA-signaling via NFKB) and pathways important for myogenesis and apical junction (**Figure 3B-C**). Only about a third of the DEGs are common to both datasets suggesting that *AFF3* LoF and DN variants largely modulate transcriptomes differently (**Figure 3A**). For example, pathways involved in the epithelium-mesenchyme transition, early response to estrogen, hypoxia, xenobiotic metabolism and apoptosis, as well as genes that are downregulated by KRAS are specifically upregulated in the KINSSHIP/KINSSHIP lines (**Figure 3B-C**). Within the set of 1272 common DEGs, 121 genes present opposite mirror effects in both strains, i.e. they are up-regulated in one genotype and downregulated in the other (**Figure 3A**). They are enriched for DNA repair genes, a pathway activated in LoF/LoF but not in KINSSHIP/KINSSHIP cells (**Figure 3C**). A core set of 20 DEGs are similarly modified upon *AFF2*, *AFF3* and *AFF4* knock-down^3^ or when *AFF3* harbors homozygous DN or LoF variants suggesting that they are sensitive to any SECs’ perturbation.

We then assessed if DEGs were direct or indirect targets of AFF3. While in excess of 3,500 genes are dysregulated in each genotype, only 226 genes presented with a neighboring AFF3 ChIP-seq peak using a FDR of 0.05%^13^ (**Table S5**), suggesting that many of the observed transcriptome changes are downstream effects. However, 32% (74 out of 226) of the bound loci were dysregulated in either the LoF/LoF and/or the DN/DN lines (**Figure 3D**). The binding sites of the orthologous mouse Aff3 were determined in ES cells by ChiP-seq^14^. Upon lifting Aff3 ChIP-seq peaks to the human genome, we similarly found that 42% of genes with a binding site (374 out of 881) were DEGs in either the LoF/LoF and/or the DN/DN lines (**Figure 3D**; **Table S6**). While such inter-clade binding comparisons have caveats, our HEK293T and ES results suggest that a substantial proportion of bound loci are dysregulated upon changes in the expression level of *AFF3* and/or stability of AFF3 (**Figure 3D-E**; **Figure S4**, **Table S7**). These dysregulated direct targets include genes associated with traits present in *AFF3* variants carriers such as neurodevelopmental disorders (e.g. *AGO1, ARV1, CDK5RAP2, CNNM2, CRADD, DPP9, EDEM3, GAN, HNRNPA2B1, HNRNPU, IRX5, MGRN1, MTRR, PREPL, SOX4*, *SRCAP*, *TOR1A TUBB, VPS35L*) and autism (i.e. part of SFARI gene list, e.g. *CDK5RAP2, CTNNA3, DAGLA, DLX3*, *LDB1, MYH10, PREX1, PRKACA, SETDB1, SRCAP* and *ST7*), ossification and limb defects (*DLX3*, *DPP9*, *DYNC2I2, FN1, IRX5,, RPS7SRCAP*, *VPS35L*), pilosity abnormalities (*DLX3*, *GAN, JUP, NR3C1*), renal diseases (*CNNM2, RPS7, ZNF423*), cardiac disorders (*CTNNA3, JUP, KIF20A, LMNA*, *VPS35L*) and dysmorphisms (*LEMD2*, *IRX5, LMNA, RPS7, SOX4*). They also comprise key genes implicated in axon guidance, cell migration and cell fate (e.g. *DDX17*, *EFNA5, FZD7*, *GAN, ISL2, JUN, MDK, MYO1D, NAV1, NOTCH2, SNAI1* and *SP1*). Importantly, some direct targets are upregulated, while others are downregulated. For example, *DDX17* is downregulated in both LoF/LoF and DN/DN lines, whereas *CTNNA3* and *NR3C1* are only downregulated in LoF/LoF lines and *CDK5RAP2* only in DN/DN lines. On the contrary, *CNNM2* is upregulated in DN/DN lines (**Figure 3E**, **Figure S4**).

As many of our affected individuals present heterozygous LoF variants, we then compared the transcriptome profiles of the LoF/+ lines with that of the LoF/LoF lines and observed that, while the same pathways are affected (**Figure S5**), only 22% of the DEGs of the homozygous line were also dysregulated in the heterozygous line, suggesting a dose dependent modification (**Table S8**). We similarly compared the transcriptomes of the compound heterozygote DN/LoF lines to those of both the LoF/LoF and DN/DN lines. While 37% of the DEGs common to LoF/LoF and DN/DN lines are modified in the DN/LoF lines, we also observe in this heterozygous line modifications in expression levels that overlap with DEGs specifically modified in either homozygous line corresponding to 22 and 27% of the DEGs specifically modified in each group of lines, respectively (**Table S9**). This suggests a co-dominance of the Ala233Thr and stop-gain variants where the increased stability of the first allele does partially compensate the decreased expression level of the second allele in some instances and over-compensate in others.

In parallel, we compared the transcriptome of primary fibroblasts from two probands with biallelic missense alterations of *AFF3* (B1: homozygous p.(Lys528Arg); B7: compound heterozygote p.(Val1036Ile)/p.(Arg1186Gln (**Figure 1A**; **Table S2**) to those of three healthy controls by RNA-sequencing. We found 142 DEGs at an adjp-value threshold of 0.05 (**Table S10**). *AFF3* mRNA levels are significantly decreased in both probands (p< 0.0002; **Table S10**) and a comparable number of distinctive reads corresponding to both alleles of the compound heterozygote were identified (**Table S11**), which is consistent with the notion that the three *AFF3* missense variants harbored by these probands are deleterious. While only nineteen and sixteen percent of the fibroblasts DEGs are also DEGs in the LoF/LoF and DN/DN HEK293T lines, respectively, the same hallmark pathways are dysregulated. For example, genes involved in the G2-M transition, targets of E2F and MYC-related genes and interferon alpha-response are enriched within their list of respective DEGs (**Table S11, Figure S5**). These results suggest that similar pathological mechanisms are at play when AFF3 is haploinsufficient and when it harbors biallelic missense variants.

## Discussion

We present evidence suggesting that multiple *AFF3* variant-specific mechanisms are associated with cognitive impairment. While somatic variants in the promoter of this gene were previously linked to mild ID^17,18^, we show that DN^11^, duplication, truncation, deletion, absence as well as biallelic variants in *AFF3* are associated with ID. Suggestive of semi-dominance, homozygous LoF (L12 and L13) and compound heterozygous LoF/missense (L10) individuals present more severe phenotypes than their heterozygous parents (**Figure 1C**). The hypothesis that non-degron *de novo* missense variants are also linked to DD/ID warrants further investigation and the identification of more affected individuals. Common variants in this locus were similarly GWAS- or MTAG-associated (multi-trait analysis of GWAS) with cognition proxies such as fluid intelligence, educational attainment, and mathematical ability, or with correlated traits such as household income, occupational attainment, and brain morphology^52^. Consistent with these findings, *AFF3* and its macaque, mouse, rat, rabbit and chicken orthologs are expressed during the early stages of brain and cerebellum development in particular in late neurons^53,54^, where it plays a direct role in the migration of cortical neurons^55^. Likewise, common variants in this locus are associated with scoliosis, anthropometric traits (BMI, height) and pulmonary involvement (vital capacity, asthma, chronic obstructive pulmonary disease), three cardinal features of KINSSHIP syndrome. *AFF3* is also GWAS-/MTAG-associated with diabetes (type 1, type 2, diabetic nephropathy and HDL cholesterol), addictions (smoking initiation, alcohol consumption, cannabis dependence, television watching), autoimmunity (lupus, celiac disease, rheumatoid and juvenile idiopathic arthritis), sexual development and dimorphism (age at menarche, endometriosis, mammographic density, male baldness, biological sex), blood measurements (e.g. hematocrit, hemoglobin measurement), eye diseases (e.g. astigmatism, intraocular pressure, corneal measurements), and insomnia^52^. This high pleiotropy is consistent with the large and diversified transcriptional role of AFF3. It suggests that any perturbation of its expression level might be deleterious. Consistent with this hypothesis we identify multiple modes-of-action and observe variant-specific/expression level modulation of the phenotype. Firstly, untimely (over)expression of KINSSHIP variants that are less sensitive to SIAH regulation leads to extremely severe phenotypes in human, zebrafish and rodents, e.g. homozygous Ala233Thr knock-in leads to mouse lethality^11^. Secondly, C57BL/6N and CD1 genetic backgrounds modulate the phenotypes presented by *Aff3* mouse knockouts^11,56,57^. Thirdly, knockdown and overexpression of mouse *Aff3* in dermal cells impair niche switching, which is required for hair reconstitution^10^. Fourthly, *AFF3* overexpression in HeLa cells perturbed the dynamics of the nuclear speckles^1^. Our RNA-seq experiments further demonstrate that changes in the amount and/or function of AFF3 dramatically alter transcriptome profiles. We show that the expression of about one-third of the AFF3 targets (bound loci) are differentially expressed upon *AFF3* modification and observe a progression in the extent of transcriptome alterations with those linked to haploinsufficiency being less drastic than that of homozygous LoF cells, which in turn are less impacted than cells harboring homozygous DN variants.

## Conclusions

In conclusion, we are adding to a growing list of variant-specific neurodevelopmental mechanisms and their associated genotype-phenotype correlations^58–60,61^. We demonstrate that beside degron variants that impair the degradation of the encoded protein^11^ and downregulation due to promoter hypermethylation^17,18^, dysregulation of *AFF3* through gene duplication, heterozygous and biallelic truncating variants, biallelic missense variants and compound heterozygous truncating/missense variants are associated with cognitive impairment.

## Data Availability

Data produced in the present study are available upon reasonable request to the authors. The HEK293T and fibroblast RNA-seq reads are deposited in GEO under accession GSE241621 (token otezqcsslbctbyx) and GSE246554 (token wxibcyqatdyphkl), and will be available from August 2024.

## List of abbreviations

ID: intellectual disabilities
DD: developmental delay
DN: dominant-negative
LoF: loss-of-function
+: wild-type allele
NHD: N-terminal homology domain
CHD: C-terminal homology domains
ALF: AF4-LAF4-FMR2 domain
TAD: transactivation domain
NLS: nuclear/nucleolar localization sequence
SECs: transcriptional super elongation complexes
P-TEF: positive transcription elongation factor
MLLT: myeloid/lymphoid or mixed-lineage leukemia; translocated to
ELL: Elongation Factor for RNA Polymerase II
ELLbow: ELL-binding domain
pLI: probability of being loss-of-function intolerant
pLOEUF: loss-of-function observed/expected upper bound fraction
PTU: 1-phenyl 2-thiourea
K22-K26: KINSSHIP patients 22 to 26
DUP: patient with partial duplication of *AFF3*
L1-L14: patients 1 to 14 with loss-of-function variants
B1-B7: patients 1 to 7 with biallelic missense variants
ADHD: attention deficit hyperactivity disorder
EEG: Electroencephalography
ASD: autism spectrum disorder
KD: knock-down
Dpf: days post fertilization
IOD: inter-ocular distance
HW: head width
Un: Untreated
M: mock
Wt: wild-type
MAF: minor allele frequency
DEG: differentially expressed genes
MTAG-associated: associated by multi-trait analysis of GWAS
BMI: body mass index

## Ethics approval, consent to participate and consent for publication

Informed consent forms were obtained for all affected individuals or their guardians participating in this study. The current study was approved by the CER (“Commission d’éthique de la recherche”) of the canton of Vaud (Protocol number: CER-VD 2021-01400). This research complies with the principles of the Declaration of Helsinki.

## Availability of data and materials

The HEK293T and fibroblast RNA-seq reads are deposited in GEO under accession GSE241621 (token otezqcsslbctbyx) and GSE246554 (token wxibcyqatdyphkl).

## Conflicts of Interests

Annabelle Tuttle, Houda Zghal Elloumi and Chaofan Zhang are employees of GeneDx and Desiree DeMille works for ARUP Laboratories. James R. Lupski has stock ownership in 23andMe and is a paid consultant for Genome International. Claudia M.B. Carvalho provides consulting service for Ionis Pharmaceuticals. The other authors have no competing interests to declare.

## Authors contributions

SB, JC, NV, BY engineered and phenotyped animal and cell models. GA, FS and CI analyzed transcriptomes profiles. NG 3D-modeled missense variants. AB, FS, LT, SS, RAJ, J-US, DD, PB-T, GRN, KNW, LD, MM, CG, LELMV, RP, RK, HY, GÅMH, CJ, MFS, KMB, MJL, CMBC, CZ, JRL, LP, LF-G, RM-T, FP, AT, HZE, LM, MK, OK, JH, MS, MI, FO, FZ, KW, AM, MKH, PP and HA collected clinical information and genomic DNAs, sequenced and analyzed exomes and/or genomes. AR and SB conceived the study and wrote the manuscript. All other authors commented on the manuscript.

## Acknowledgments and Fundings

We thank Jacques Beckmann for comments. This work was supported by grants from the Swiss National Science Foundation (31003A_182632 and IZSTZ0_216615 to AR), the Lejeune Foundation (#1838-2019A to AR), the Blackswan Foundation (to AR), a PRIN 2020 grant from the Italian Ministry of Universities and Research (20203P8C3X to AB), and the US National Institutes of Health (NS105078 and HG011758 to JRL). This study makes use of data generated by the DECIPHER community. Funding for the DECIPHER project was provided by the Wellcome Trust [grant number WT223718/Z/21/Z]. The funders had no role in study design, data collection and analysis, decision to publish, or preparation of the manuscript.

## Supplementary Figures

**Supplementary Figure S1.**
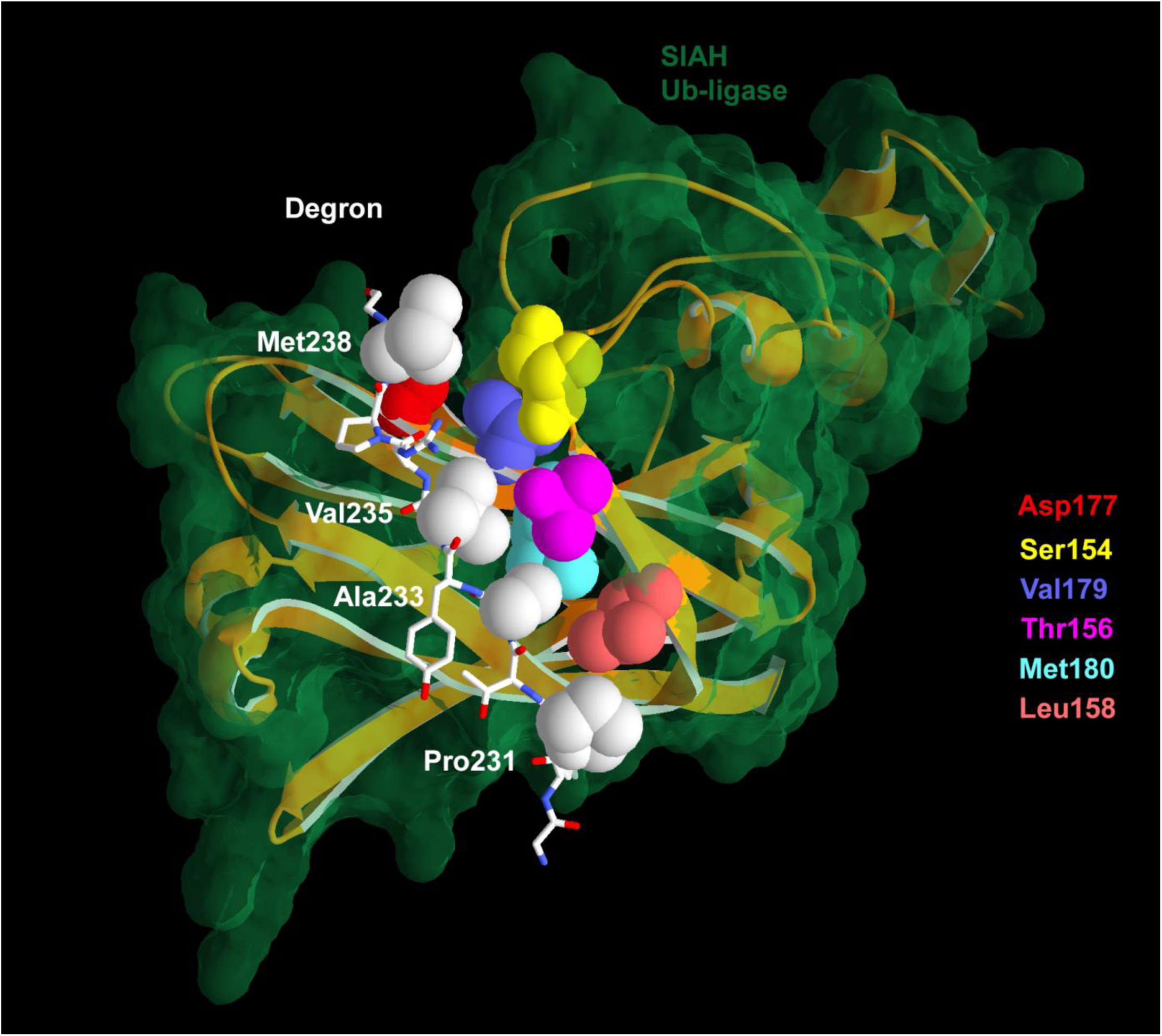
3D protein modelling of the human AFF3 degron region bound to SIAH ubiquitin ligase. The AFF3 degron chain is shown as a white stick structure, with residues mutated in KINSSHIP affected individuals highlighted in white space-fill, from top to bottom: Met238, Val235, Ala233, and Pro231. The SIAH ubiquitin ligase is presented as an orange ribbon embedded in its green transparent surface. Amino acids interacting with the degron residues sidechains are clustered in two spatial regions along beta strands and represented space-filled as follow: Ser154 yellow, Thr156 burgundy Leu158 salmon and Asp177 red, Val179 blue, Met180 cyan.

**Supplementary Figure S2.**
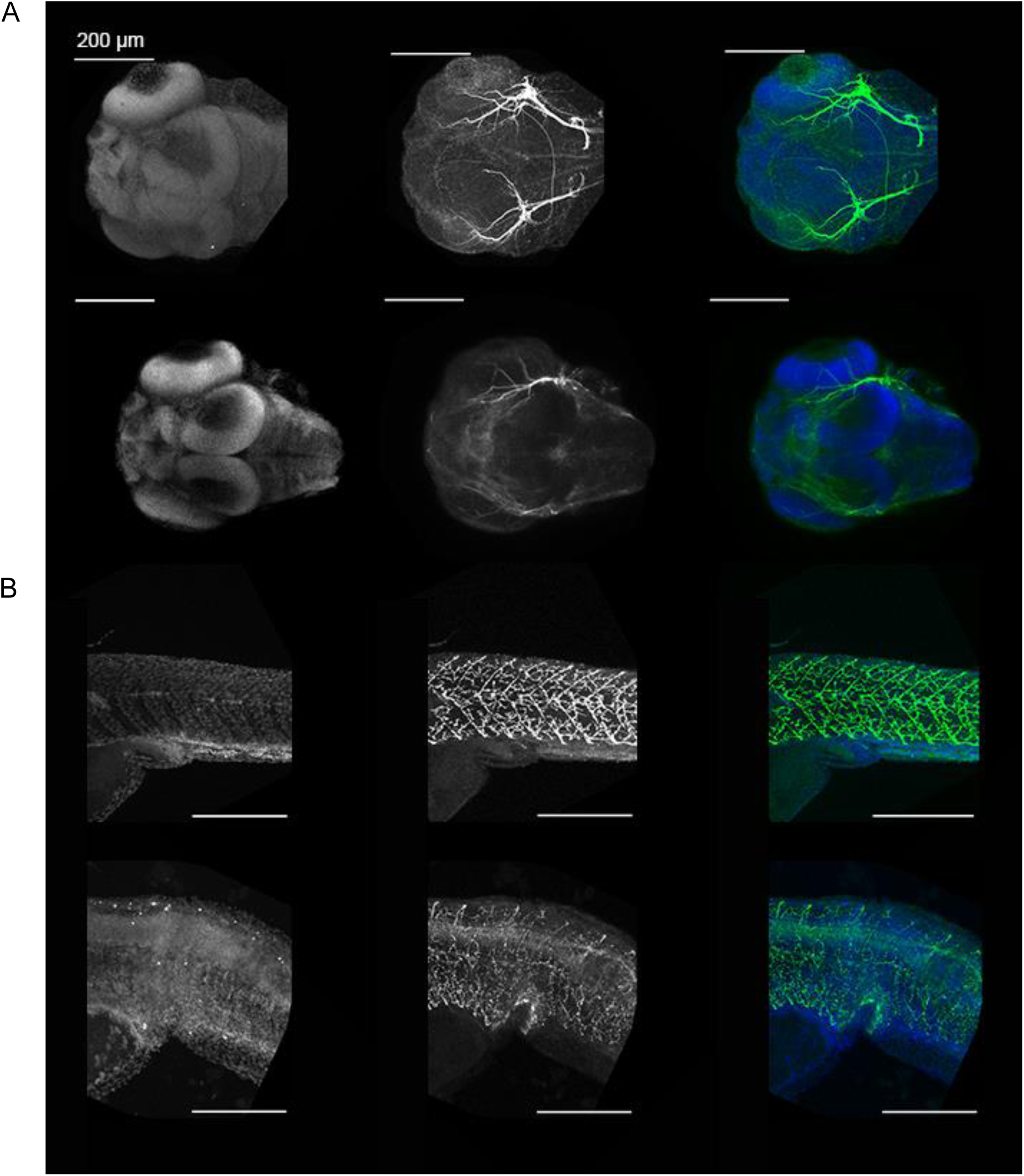
Immunostaining of hindbrain neurons and motoneurons in 3dpf zebrafish. Maximum projections of confocal images regarding hindbrain neuronal structures (A) and motoneurons (B) in 3dpf Mock-injected (Mock) and *aff3* KD larvae.

**Supplementary Figure S3.**
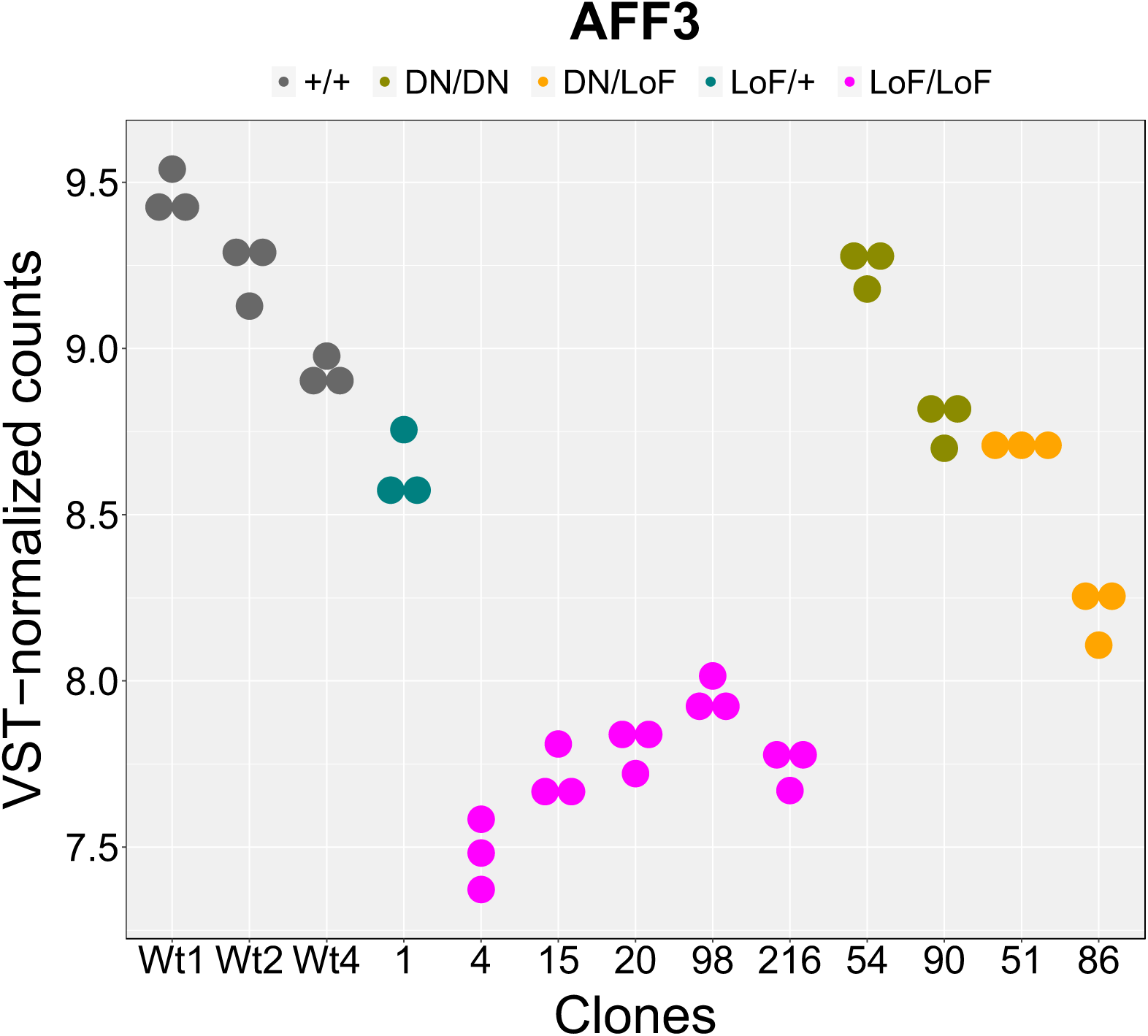
AFF3 expression levels in engineered isogenic HEK293T cells. Dotblot of the VST-normalized (r-log) counts of the *AFF3* gene across all HEK293T engineered samples. We engineered five biallelic LoF HEK293T lines (LoF/LoF) with different combinations of variants (lines No.20 and 98: stop-gain/stop-gain; No.15: stop-gain/20bp deletion; No.4: 4bp deletion/114bp deletion; No.216: 94bp deletion/94bp deletion), one heterozygous LoF stop-gain/+ line (No.1), two homozygous Ala233Thr/Ala233Thr KINSSHIP/KINSSHIP lines (No.54 and 90) and two compound heterozygous KINSSHIP and LoF lines (No.51 and 86: Ala233Thr/stop-gain). The variant nomenclature of the engineered variants is specified in the materials and methods section.

**Supplementary Figure S4.**
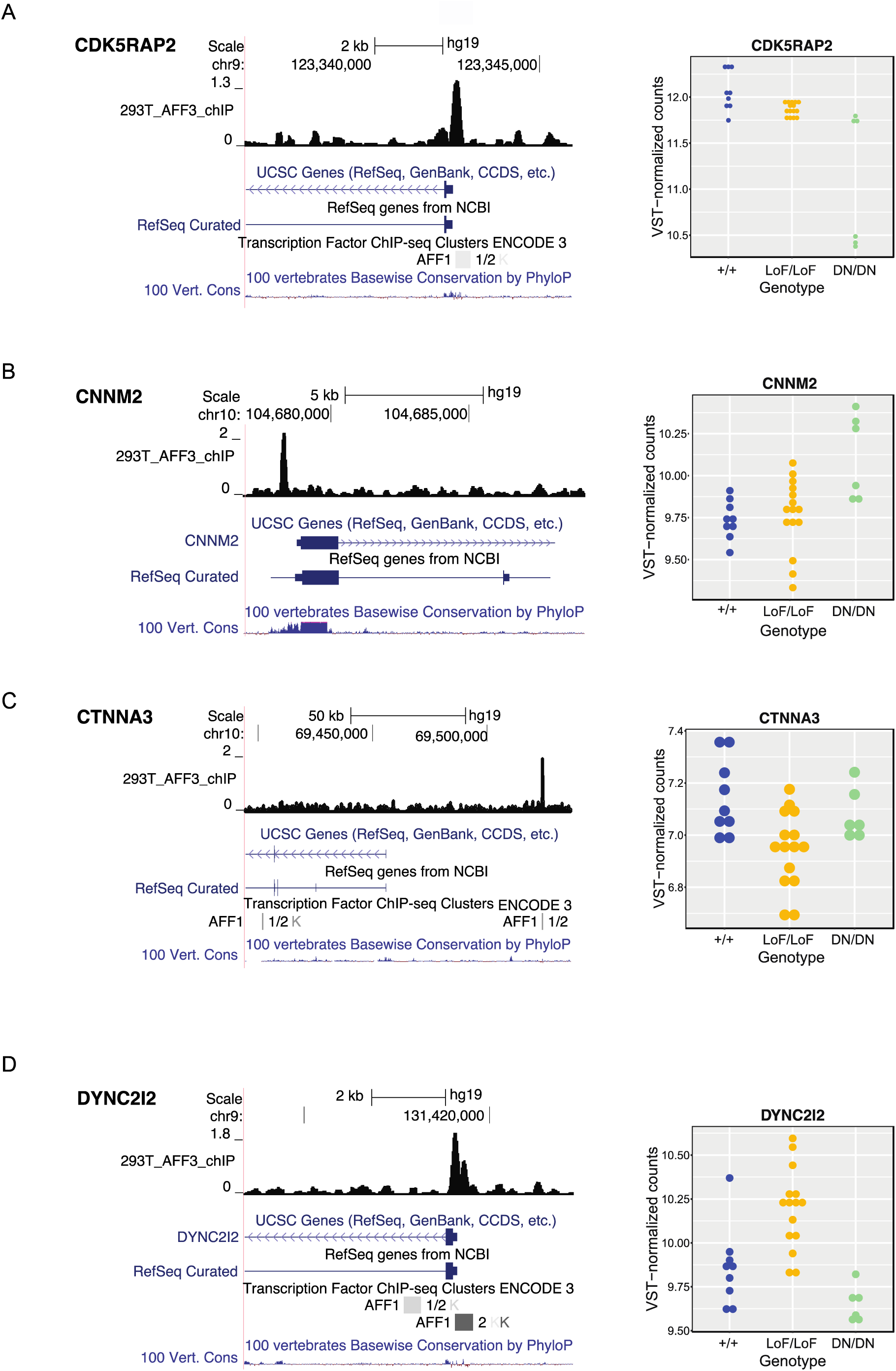
Examples of DEGs loci bound by AFF3. UCSC genome browser snapshot of the CDK5RAP2, CNNM2, CTNNA3 and DYN*C2I2* loci a bound by AFF3 showing from to top to bottom AFF3 ChIP-seq HEK293T results, UCSC and REFSeq curated gene structure and vertebrate PhyloP conservation scores (left panels). Expression level of the corresponding DEGs in +/+ (blue), LoF/LoF (yellow) and DN/DN (green) HEK293T engineered lines (right panels).

**Supplementary Figure S5.**
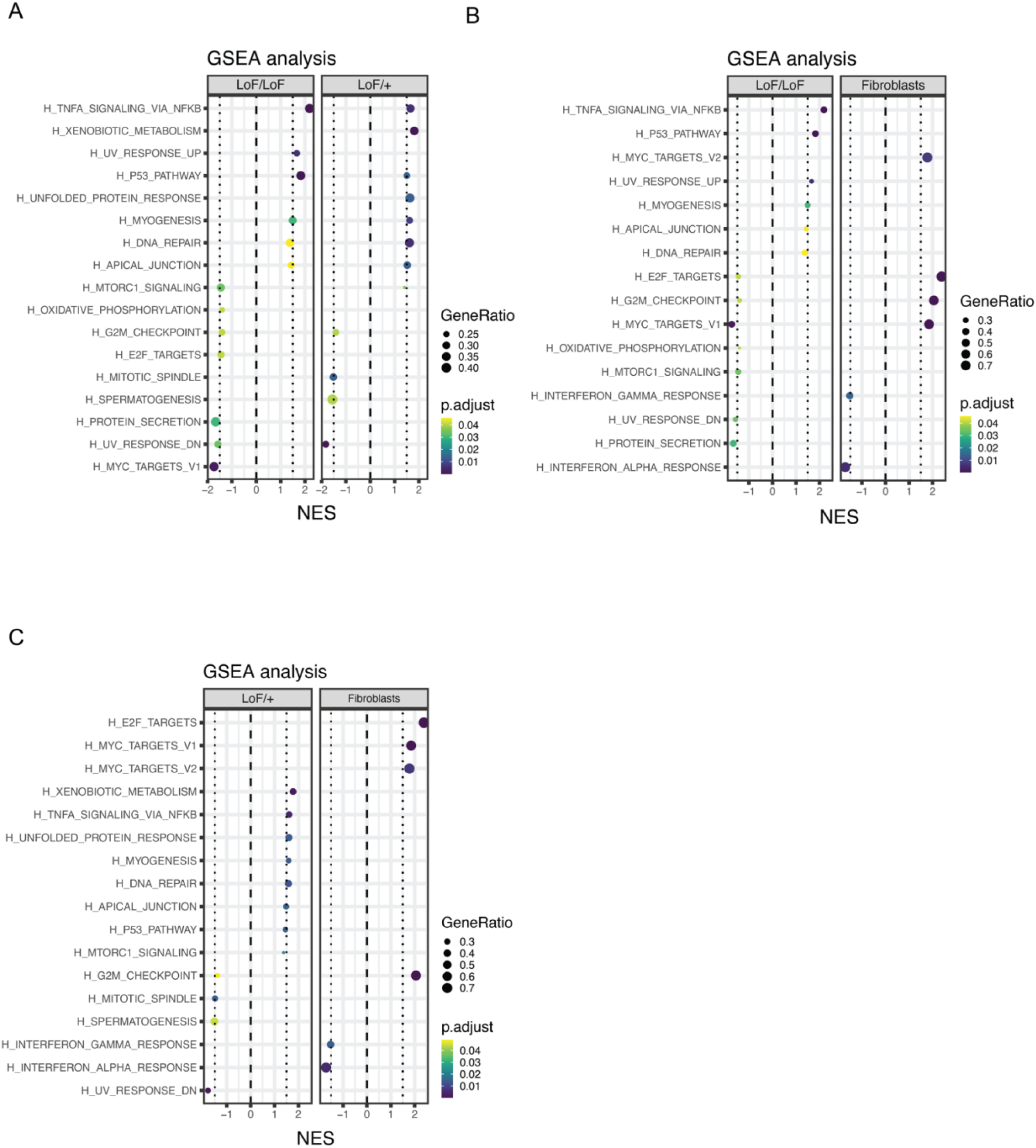
Gene Set Enrichment Analysis. Gene Set Enrichment Analysis (GSEA) for hallmark pathways of DEGs **(A)** in biallelic loss-of function (LoF/LoF) *AFF3* lines (left panel) and heterozygote loss-of function (Lof/+) *AFF3* lines (right panel) upon comparison with unmutated wildtype lines; **(B)** in biallelic loss-of function (LoF/LoF) *AFF3* lines (left panel) and fibroblasts of probands (fibroblasts; right panel) upon comparison with unmutated wildtype lines and fibroblasts of controls, respectively; and **(C)** in heterozygous loss-of function (LoF/+) *AFF3* lines (left panel) and fibroblasts of probands (fibroblasts; right panel) upon comparison with unmutated wildtype lines and fibroblasts of controls, respectively

## Notes

### Author Declarations

Informed consent forms were obtained for all affected individuals or their guardians participating in this study. The current study was approved by the CER (Commission d'éthique de la recherche) of the canton of Vaud (Protocol number: CER-VD 2021-01400). This research complies with the principles of the Declaration of Helsinki.

## Bibliography

1. Melko, M. et al. Functional characterization of the AFF (AF4/FMR2) family of RNA-binding proteins: insights into the molecular pathology of FRAXE intellectual disability. Hum Mol Genet 20, 1873–85 (2011).

2. Guo, C. et al. The super elongation complex (SEC) mediates phase transition of SPT5 during transcriptional pause release. EMBO Rep 24, e55699 (2023).

3. Luo, Z. et al. The super elongation complex family of RNA polymerase II elongation factors: gene target specificity and transcriptional output. Mol Cell Biol 32, 2608–17 (2012).

4. Chen, Y. & Cramer, P. Structure of the super-elongation complex subunit AFF4 C-terminal homology domain reveals requirements for AFF homo- and heterodimerization. J Biol Chem 294, 10663–10673 (2019).

5. Nilson, I. et al. Exon/intron structure of the human AF-4 gene, a member of the AF-4/LAF-4/FMR-2 gene family coding for a nuclear protein with structural alterations in acute leukaemia. Br J Haematol 98, 157–69 (1997).

6. House, C.M. et al. A binding motif for Siah ubiquitin ligase. Proc Natl Acad Sci U S A 100, 3101–6 (2003).

7. Jonkers, I. & Lis, J.T. Getting up to speed with transcription elongation by RNA polymerase II. Nat Rev Mol Cell Biol 16, 167–77 (2015).

8. Wang, Y. et al. A permissive chromatin state regulated by ZFP281-AFF3 in controlling the imprinted Meg3 polycistron. Nucleic Acids Res 45, 1177–1185 (2017).

9. Tsukumo, S.I. et al. AFF3, a susceptibility factor for autoimmune diseases, is a molecular facilitator of immunoglobulin class switch recombination. Sci Adv 8, eabq0008 (2022).

10. Takeo, M. et al. Cyclical dermal micro-niche switching governs the morphological infradian rhythm of mouse zigzag hair. Nat Commun 14, 4478 (2023).

11. Voisin, N. et al. Variants in the degron of AFF3 are associated with intellectual disability, mesomelic dysplasia, horseshoe kidney, and epileptic encephalopathy. Am J Hum Genet 108, 857–873 (2021).

12. Veitia, R.A. AFF3: a new player in maintaining XIST monoallelic expression. J Mol Cell Biol 11, 723–724 (2019).

13. Zhang, Y. et al. AFF3-DNA methylation interplay in maintaining the mono-allelic expression pattern of XIST in terminally differentiated cells. J Mol Cell Biol 11, 761–769 (2019).

14. Luo, Z. et al. Regulation of the imprinted Dlk1-Dio3 locus by allele-specific enhancer activity. Genes Dev 30, 92–101 (2016).

15. Inoue, Y. et al. Three KINSSHIP syndrome patients with mosaic and germline AFF3 variants. Clin Genet 103, 590–595 (2023).

16. Karczewski, K.J. et al. The mutational constraint spectrum quantified from variation in 141,456 humans. Nature 581, 434–443 (2020).

17. Jadhav, B., et al. A GCC repeat expansion in AFF3 is a significant cause of intellectual disability. medRxiv (2023).

18. Metsu, S. et al. FRA2A is a CGG repeat expansion associated with silencing of AFF3. PLoS Genet 10, e1004242 (2014).

19. Mattioli, F. et al. Biallelic truncation variants in ATP9A are associated with a novel autosomal recessive neurodevelopmental disorder. NPJ Genom Med 6, 94 (2021).

20. Alfaiz, A.A. et al. TBC1D7 mutations are associated with intellectual disability, macrocrania, patellar dislocation, and celiac disease. Hum Mutat 35, 447–51 (2014).

21. Goujon, M. et al. A new bioinformatics analysis tools framework at EMBL-EBI. Nucleic Acids Res 38, W695–9 (2010).

22. Sievers, F. et al. Fast, scalable generation of high-quality protein multiple sequence alignments using Clustal Omega. Mol Syst Biol 7, 539 (2011).

23. Guex, N. & Peitsch, M.C. SWISS-MODEL and the Swiss-PdbViewer: an environment for comparative protein modeling. Electrophoresis 18, 2714–23 (1997).

24. Labun, K. et al. CHOPCHOP v3: expanding the CRISPR web toolbox beyond genome editing. Nucleic Acids Res 47, W171–W174 (2019).

25. Montague, T.G., Cruz, J.M., Gagnon, J.A., Church, G.M. & Valen, E. CHOPCHOP: a CRISPR/Cas9 and TALEN web tool for genome editing. Nucleic Acids Res 42, W401–7 (2014).

26. Bassani, S. et al. Variants in USP48 encoding ubiquitin hydrolase are associated with autosomal dominant non-syndromic hereditary hearing loss. Hum Mol Genet 30, 1785–1796 (2021).

27. Schindelin, J., et al. Fiji: an open-source platform for biological-image analysis. Nat Methods 9, 676–82 (2012).

28. Dobin, A. et al. STAR: ultrafast universal RNA-seq aligner. Bioinformatics 29, 15–21 (2013).

29. Liao, Y., Smyth, G.K. & Shi, W. featureCounts: an efficient general purpose program for assigning sequence reads to genomic features. Bioinformatics 30, 923–30 (2014).

30. Love, M.I., Huber, W. & Anders, S. Moderated estimation of fold change and dispersion for RNA-seq data with DESeq2. Genome Biol 15, 550 (2014).

31. Huber, W. et al. Orchestrating high-throughput genomic analysis with Bioconductor. Nat Methods 12, 115–21 (2015).

32. Wu, T. et al. clusterProfiler 4.0: A universal enrichment tool for interpreting omics data. Innovation (Camb) 2, 100141 (2021).

33. Yu, G., Wang, L.G., Han, Y. & He, Q.Y. clusterProfiler: an R package for comparing biological themes among gene clusters. OMICS 16, 284–7 (2012).

34. Subramanian, A. et al. Gene set enrichment analysis: a knowledge-based approach for interpreting genome-wide expression profiles. Proc Natl Acad Sci U S A 102, 15545–50 (2005).

35. Liberzon, A. et al. Molecular signatures database (MSigDB) 3.0. Bioinformatics 27, 1739–40 (2011).

36. Liberzon, A. et al. The Molecular Signatures Database (MSigDB) hallmark gene set collection. Cell Syst 1, 417–425 (2015).

37. Zhang, Y. et al. Model-based analysis of ChIP-Seq (MACS). Genome Biol 9, R137 (2008).

38. Yu, G., Wang, L.G. & He, Q.Y. ChIPseeker: an R/Bioconductor package for ChIP peak annotation, comparison and visualization. Bioinformatics 31, 2382–3 (2015).

39. Deciphering Developmental Disorders, S. Prevalence and architecture of de novo mutations in developmental disorders. Nature 542, 433–438 (2017).

40. Sobreira, N., Schiettecatte, F., Valle, D. & Hamosh, A. GeneMatcher: a matching tool for connecting investigators with an interest in the same gene. Hum Mutat 36, 928–30 (2015).

41. Firth, H.V. et al. DECIPHER: Database of Chromosomal Imbalance and Phenotype in Humans Using Ensembl Resources. Am J Hum Genet 84, 524–33 (2009).

42. Jacquemont, S. et al. A higher mutational burden in females supports a “female protective model” in neurodevelopmental disorders. Am J Hum Genet 94, 415–25 (2014).

43. Harripaul, R. et al. Mapping autosomal recessive intellectual disability: combined microarray and exome sequencing identifies 26 novel candidate genes in 192 consanguineous families. Mol Psychiatry 23, 973–984 (2018).

44. Tang, D. et al. Structural and functional insight into the effect of AFF4 dimerization on activation of HIV-1 proviral transcription. Cell Discov 6, 7 (2020).

45. Qi, S. et al. Structural basis for ELL2 and AFF4 activation of HIV-1 proviral transcription. Nat Commun 8, 14076 (2017).

46. Kalueff, A.V. et al. Towards a comprehensive catalog of zebrafish behavior 1.0 and beyond. Zebrafish 10, 70–86 (2013).

47. Sillar, K.T. Mauthner cells. Curr Biol 19, R353–5 (2009).

48. Skarnes, W.C. et al. A conditional knockout resource for the genome-wide study of mouse gene function. Nature 474, 337–42 (2011).

49. Khan, H. et al. A novel variant in AFF3 underlying isolated syndactyly. Clin Genet 103, 341–345 (2023).

50. Niederriter, A.R. et al. In vivo modeling of the morbid human genome using Danio rerio. J Vis Exp, e50338 (2013).

51. Ioannidis, N.M. et al. REVEL: An Ensemble Method for Predicting the Pathogenicity of Rare Missense Variants. Am J Hum Genet 99, 877–885 (2016).

52. Sollis, E. et al. The NHGRI-EBI GWAS Catalog: knowledgebase and deposition resource. Nucleic Acids Res 51, D977–D985 (2023).

53. Telley, L. et al. Sequential transcriptional waves direct the differentiation of newborn neurons in the mouse neocortex. Science 351, 1443–6 (2016).

54. Cardoso-Moreira, M. et al. Gene expression across mammalian organ development. Nature 571, 505–509 (2019).

55. Moore, J.M. et al. Laf4/Aff3, a gene involved in intellectual disability, is required for cellular migration in the mouse cerebral cortex. PLoS One 9, e105933 (2014).

56. Kraft, K. et al. Deletions, Inversions, Duplications: Engineering of Structural Variants using CRISPR/Cas in Mice. Cell Rep 10, 833–839 (2015).

57. Birling, M.C. et al. A resource of targeted mutant mouse lines for 5,061 genes. Nat Genet 53, 416–419 (2021).

58. den Hoed, J., et al. Mutation-specific pathophysiological mechanisms define different neurodevelopmental disorders associated with SATB1 dysfunction. Am J Hum Genet 108, 346–356 (2021).

59. O’Donnell-Luria, A.H. et al. Heterozygous Variants in KMT2E Cause a Spectrum of Neurodevelopmental Disorders and Epilepsy. Am J Hum Genet 104, 1210–1222 (2019).

60. Haijes, H.A. et al. De Novo Heterozygous POLR2A Variants Cause a Neurodevelopmental Syndrome with Profound Infantile-Onset Hypotonia. Am J Hum Genet 105, 283–301 (2019).

61. Paul, M.S. et al. Rare EIF4A2 variants are associated with a neurodevelopmental disorder characterized by intellectual disability, hypotonia, and epilepsy. Am J Hum Genet 110, 548 (2023).

