## Supplemental Table S1 for "Variant-specific pathophysiological mechanisms of *AFF3* differently influence transcriptome profiles"

| **Table S1: genotype of KINSSHIP individuals** | | | | | |  |  |
| --- | --- | --- | --- | --- | --- | --- | --- |
| **ID patient** | K1 - K19, Voisin et al. AJHG 2021 (for comparison) | K22 | K23 | K24 | K25, DDD study, Nature 2017, sample DDD4K.02548 | K26, DDD study, Nature 2017, sample DDD4K.00047 | DUP1 |
| **Variant in AFF3**  **Ref ENST00000317233.4** |  | GRCh37:2:100623254:A>G, NM_002285.3:c.713T>C, NP_002276.2:p.(Met238Thr); | GRCh37:2:100623276:G>A, NM_002285.3:c.691C>T, NP_002276.2:p.(Pro231Ser); | GRCh37:2:100623270:C>T  NM_002285.3: c.697G>A,  NP_002276.2: p.(Ala233Thr) | GRCh37:2:100623270:C>T  NM_002285.3: c.697G>A,  NP_002276.2: p.(Ala233Thr) | GRCh37:2:100623270:C>A  NM_002285.3: c.697G>T,  NP_002276.2: p.(Ala233Ser) | chr2:g.100077649_100359928dup (hg19) (NC_000002.11:g.100077649_100359928dup) |
| **GnomAD** | All not reported | Not reported | Not reported | Not reported | Not reported | Not reported | Not reported |
| **Prediction tools** |  | SIFT = 0, POLYPHEN = 0.99, CADD = 23.6, REVEL = 0.623 | SIFT = 0, POLYPHEN = 0.99, CADD = 25.4, REVEL = 0.769 | CADD=27.1, REVEL = 0.608 | CADD=27.1, REVEL = 0.608 |  |  |
| **Status of the variant** | Heterozygote | Heterozygote | Heterozygote | Heterozygote | Heterozygote | Heterozygote | Heterozygote |
| **Inheritance** | All *de novo* | *de novo* | *de novo* | *de novo* | *de novo* | *de novo* | *de novo* |
