## Supplemental Table S2 for "Variant-specific pathophysiological mechanisms of *AFF3* differently influence transcriptome profiles"

| **Table S2: genotype of carriers of AFF3 variants** |  |  |  |  |  |  |  |  |  |  |  |  |  |  |  |  |  |  |  |  |  |  |  |  |  |
| --- | --- | --- | --- | --- | --- | --- | --- | --- | --- | --- | --- | --- | --- | --- | --- | --- | --- | --- | --- | --- | --- | --- | --- | --- | --- |
| **ID Patient** | L1 | L2 | L3 | L4 | L5 | L6 | L7  DECIPHER sample 281982 | L8  DECIPHER  Sample 305794 | L9 | L10 | L11 | L12 | L13 | L14 |  | B1 | B2 | B3 | B4  Harripaul, Mol Psy 2018, family PK113 | B5  Harripaul, Mol Psy 2018, family PK113 | B6  Harripaul, Mol Psy 2018, family PK113 | B7 |  | M1 | M2  DDD study, Nature 2017, sample DDD4K.02548 |
| **Family** | Family 1 | Family 1 | Family 1 | Family 1 | Family 2 | Family 3 | Family 4 | Family 5 | Family 6 | Family 6 | Family 7 | Family 8 | Family 8 | Family 8 |  | Family 9 | Family 10 | Family 10 | Family 11 | Family 11 | Family 11 | Family 12 |  | Family 13 | Family 14 |
| **Family member** | Proband | Brother 1 | Brother 2 | Father | Proband | Proband | Proband | Proband | Father | Proband | Proband | Proband | Brother | Mother |  | Proband | Proband | Sister | Affected 1 | Affected 2 | Affected 3 | Proband |  | Proband | Proband |
| **Variant 1**  **Ref ENST00000317233.4** | GRCh37:2:100170944:CGGAGCTGGCG>C; NM_002285.3:c.3378_3387delGGAGCTGGCG, NP_002276.2:p.(Ala1127TrpfsTer134) cDNA.3614_3623delCGCCAGCTCC | GRCh37:2:100170944:CGGAGCTGGCG>C; NM_002285.3:c.3378_3387delGGAGCTGGCG, NP_002276.2:p.(Ala1127TrpfsTer134) cDNA.3614_3623delCGCCAGCTCC | GRCh37:2:100170944:CGGAGCTGGCG>C; NM_002285.3:c.3378_3387delGGAGCTGGCG, NP_002276.2:p.(Ala1127TrpfsTer134) cDNA.3614_3623delCGCCAGCTCC | GRCh37:2:100170944:CGGAGCTGGCG>C; NM_002285.3:c.3378_3387delGGAGCTGGCG, NP_002276.2:p.(Ala1127TrpfsTer134) cDNA.3614_3623delCGCCAGCTCC | 570kb deletion of 2q11.2 (GRCh37,NC_000002.11:g.100456588_101026865del) | GRCh37:2:100199329:A>AAACAAACTGTTGCC, NM_002285.3:c.2710_2723dup, NP_002276.2:p.(Phe908LeufsTer85) | arr[GRCh38] 2q11.2(99644464_100282475)x1 pat(GRCh37_100260927_100898938del) | 744.09 kb deletion, GRCh37:2:99487537_100232437del, NC_000002.11:g.99487537_100232437del) | GRCh37:2:100209897G>GA, NM_002285.3:c.2225dup, NP_002276.2:p.(Tyr743LeufsTer19) | GRCh37:2:100209897G>GA, NM_002285.3:c.2225dup, NP_002276.2:p.(Tyr743LeufsTer19) | GRCh37:2:100623679:GC>G, NM_002285.3:c.417del, NP_002276.2:p.(Gln139HisfsTer82) | GRCh37:2:100368748:GA>G, NM_002285.3:c.936del, NP_002276.2:p.(Pro313HisfsTer60) | GRCh37:2:100368748:GA>G, NM_002285.3:c.936del, NP_002276.2:p.(Pro313HisfsTer60) | GRCh37:2:100368748:GA>G, NM_002285.3:c.936del, NP_002276.2:p.(Pro313HisfsTer60) |  | GRCh37:2:100210540:T>C, NM_002285.3:c.1583A>G, NP_002276.2:p.(Lys528Arg) | GRCh37:2:100210342:G:C, NM_002285.3:c.1781C>G, NP_002276.2:p.(Thr594Ser) | GRCh37:2:100210342:G>C, NM_002285.3:c.1781C>G, NP_002276.2:p.(Thr594Ser) | GRCh37:2:100167973:C>A, NM_002285.3:c.3644G>T, NP_002276.2:p.(Gly1215Val) | GRCh37:2:100167973:C>A, NM_002285.3:c.3644G>T, NP_002276.2:p.(Gly1215Val) | GRCh37:2:100167973:C>A, NM_002285.3:c.3644G>T, NP_002276.2:p.(Gly1215Val) | GRCh37:2: 100181962:C>T, NM_002285.3: c.3106G>A, NP_002276.2: p.(Val1036Ile) |  | GRCh37:2:100199397:C:T, NM_002285.3:c.2656G>A, NP_002276.2:p.(Ala886Thr) | GRCh37:2: 100368751 G>A, NM_002285.3:c.934C>T,  NP_002276.2:p.(Leu312Phe) |
| **Variant 1 in Gnomad** | Not reported | Not reported | Not reported | Not reported | Not reported | Not reported | Not reported | Not reported | Not reported | Not reported | Not reported | Not reported | Not reported | Not reported |  | Not reported | Not reported | Not reported | Not reported | Not reported | Not reported | 4E-05 |  | Not reported | Not reported |
| **Prediction tools** |  |  |  |  |  |  |  |  | SpliceAI = 0.99 (splice loss) | SpliceAI = 0.99 (splice loss) | Splice AI = 0.22 (splice loss) |  |  |  |  | SIFT = 0.45, POLYPHEN = 0.029, CADD = 15.57, REVEL = 0.044 | SIFT = 0.09, POLYPHEN = 0.038, CADD = 18.49, REVEL = 0.172 | SIFT = 0.09, POLYPHEN = 0.038, CADD = 18.49, REVEL = 0.172 | SIFT = 1, POLYPHEN = 0.98, CADD = 26, REVEL = 0.723 | SIFT = 1, POLYPHEN = 0.98, CADD = 26, REVEL = 0.723 | SIFT = 1, POLYPHEN = 0.98, CADD = 26, REVEL = 0.723 | SIFT = 0, POLYPHEN = 0.774, CADD = 27.4, REVEL = 0.298 |  | SIFT = 0.006, POLYPHEN = 0.003, CADD = 16.72, REVEL = 0.044 | SIFT = 0.16, POLYPHEN = 0.785, CADD = 24.6, REVEL = 0.247 |
| **Variant 2**  **Ref ENST00000317233.4** | Does not apply | Does not apply | Does not apply | Does not apply | Does not apply | Does not apply | Does not apply | Does not apply | Does not apply | GRCh37:2:100182009:T>C, NM_002285.3:c.3059A>G, NP_002276.2:p.(Gln1020Arg) | GRCh37:2:100623679:GC>G, NM_002285.3:c.417del, NP_002276.2:p.(Gln139HisfsTer82) | GRCh37:2:100368748:GA>G, NM_002285.3:c.936del, NP_002276.2:p.(Pro313HisfsTer60) | GRCh37:2:100368748:GA>G, NM_002285.3:c.936del, NP_002276.2:p.(Pro313HisfsTer60) | Does not apply |  | GRCh37:2:100210540:T>C, NM_002285.3:c.1583A>G, NP_002276.2:p.(Lys528Arg) | GRCh37:2:100210342:G:C, NM_002285.3:c.1781C>G, NP_002276.2:p.(Thr594Ser) | GRCh37:2:100210342:G>C, NM_002285.3:c.1781C>G, NP_002276.2:p.(Thr594Ser) | GRCh37:2:100167973:C>A, NM_002285.3:c.3644G>T, NP_002276.2:p.(Gly1215Val) | GRCh37:2:100167973:C>A, NM_002285.3:c.3644G>T, NP_002276.2:p.(Gly1215Val) | GRCh37:2:100167973:C>A, NM_002285.3:c.3644G>T, NP_002276.2:p.(Gly1215Val) | GRCh37:2:100170775:C>T, NM_002285.3:c.3557G>A, NP_002276.2:p.(Arg1186Gln) |  | Does not apply | Does not apply |
| **Variant 2 in Gnomad** |  |  |  |  |  |  |  |  |  | Not reported | Not reported | Not reported | Not reported |  |  | Not reported | Not reported | Not reported | Not reported | Not reported | Not reported | 1E-03* |  |  |  |
| **Prediction tools** |  |  |  |  |  |  |  |  |  | SIFT = 0.35, Polyphen =0.08, CADD = 19.04 , REVEL = 0.055 | Splice AI = 0.22 (splice loss) |  |  |  |  | SIFT = 0.45, POLYPHEN = 0.029, CADD = 15.57, REVEL = 0.044 | SIFT = 0.09, POLYPHEN = 0.038, CADD = 18.49, REVEL = 0.172 | SIFT = 0.09, POLYPHEN = 0.038, CADD = 18.49, REVEL = 0.172 | SIFT = 1, POLYPHEN = 0.98, CADD = 26, REVEL = 0.723 | SIFT = 1, POLYPHEN = 0.98, CADD = 26, REVEL = 0.723 | SIFT = 1, POLYPHEN = 0.98, CADD = 26, REVEL = 0.723 | SIFT = 0.2, POLYPHEN = 0.04, CADD = 24.26, REVEL = 0.150 |  |  |  |
| **Hetero/Homozygote** | Heterozygote LoF/+ | Heterozygote LoF/+ | Heterozygote LoF/+ | Heterozygote LoF/+ | Heterozygote LoF/+ | Heterozygote LoF/+ | Heterozygote LoF/+ | Heterozygote LoF/+ | Heterozygote LoF/+ | Compound heterozygote LoF/missense | Homozygote LoF/LoF | Homozygote LoF/LoF | Homozygote LoF/LoF | Heterozygote LoF/+ |  | Homozygote missense/missense | Homozygote missense/missense | Homozygote missense/missense | Homozygote missense/missense | Homozygote missense/missense | Homozygote missense/missense | Compound heterozygote missense/missense |  | Heterozygote missense/+ | Heterozygote missense/+ |
| **Inheritance** | AD, haploinsufficiency | AD, haploinsufficiency | AD, haploinsufficiency | AD, haploinsufficiency | AD, **of note, parental testing not completed** | AD, haploinsufficiency | Paternally inherited, AD, haploinsufficiency | AD, haploinsufficiency | AD, haploinsufficiency | AR (Semi-dominant ?) | AR | AR (Semi-dominant ?) | AR (Semi-dominant ?) | AD, haploinsufficiency |  | AR | AR | AR | AR | AR | AR | AR |  | AD,  *de novo* | AD,  *de novo* |
| **Sex** | M | M | M | M | M | F | F | M | M | M | M | F | M | F |  | F | F | F |  |  |  | M |  | M | F |

Footnotes: *We identified the p.(Arg1186Gln) variant in 260 genotyped alleles out of a total of 524,644 alleles investigated or a frequency of 4.96E-04 (similar to the frequency found by GnomAD) with two homozygous individuals marked as unaffected but not thoroughly phenotyped. Of note this allele is associated with a significant decrease in *AFF3* expression in fibroblasts (see text for details).
